# Safety and Efficacy of Oral administrated Cepharanthine in Non-hospitalized, asymptomatic or mild COVID-19 patients: A Double-blind, Randomized, Placebo-controlled Trial

**DOI:** 10.1101/2023.01.11.23284098

**Authors:** Jianyi Wei, Shuming Pan, Shupeng Liu, Biyun Qian, Zixuan Shen, Yan Zhang, Yuexiang Bian, ADila ABuduaini, Fuchen Dong, Xin Zhang, Jinhui Li, Yongpei Yu, Weituo Zhang, Jun Wang, Wei Zhai, Qixiang Song, Yu Zheng, Lei Li, Weihua Pan, Lanlan Yu, Qimin Zhan, Ning Zhang, Junhua Zheng, Chen Yao, Hai Li

**Affiliations:** Department of Gastroenterology, Renji Hospital, Shanghai Jiao Tong University School of Medicine; NHC Key Laboratory of Digestive Diseases (Renji Hospital, Shanghai Jiaotong University School of Medicine), 1630 Dong Fang Road, 200127, Shanghai, China; Department of Emergency, Xinhua Hospital, Shanghai Jiao Tong University School of Medicine, Yangpu District, Shanghai, China; Peking University Clinical Research Institute, Peking University First Hospital, Beijing, China; Clinical Research Center, Shanghai Jiao Tong University School of Medicine, Shanghai, China; Department of Interventional Oncology, Renji Hospital, Shanghai Jiao Tong University School of Medicine, No. 160 Pujian Rd, Pudong, Shanghai 200127, China; Department of Urology, Renji Hospital, Shanghai Jiao Tong University School of Medicine; Department of Respiratory Medicine, Renji Hospital, Shanghai Jiao Tong University School of Medicine, Shanghai, China; Department of Otorhinolaryngology-Head & Neck Surgery, Xinhua Hospital, Shanghai Jiao Tong University School of Medicine, Shanghai, China; Peking University - Yunnan Baiyao International Medical Research Center, Beijing, China; Hongqiao International Institute of Medicine, Shanghai Tongren Hospital/School of Public Health, Shanghai Jiao Tong University School of Medicine; Department of Pediatric Surgery, Xinhua Hospital, Shanghai Jiao Tong University School of Medicine, Shanghai, China

**Author notes:** **Corresponding authors:** Hai Li, Chen Yao, **Hai Li**, Address: Department of Gastroenterology, Ren Ji Hospital, Shanghai Jiao Tong University School of Medicine; NHC Key Laboratory of Digestive Diseases (Renji Hospital, Shanghai Jiaotong University School of Medicine), 1630 Dong Fang Road, 200127, Shanghai, China, Or, **Chen Yao**, Address: Peking University Clinical Research Institute, Peking University First Hospital, 100083, Beijing, China.

## Abstract

Cepharanthine (CEP) is a natural remedy that potently inhibits SARS-CoV-2 activity both in vitro and in vivo. We conducted a proof-of-concept, double-blind, randomized, placebo-controlled trial among adults with asymptomatic or mild coronavirus disease 2019 (COVID-19). Patients were stratified randomly to de novo infection or viral rebound, and assigned in a 1:1:1 ratio to receive 60 mg/day or 120 mg/day of CEP or placebo. Primary outcome the time from randomization to negative nasopharyngeal swab, and safety were evaluated. A total of 262 de novo infected and 124 viral rebound patients underwent randomization. In the 188 de novo patients included in modified intention-to-treat (mITT) population, when compared with placebo, 60 mg/day CEP slightly shortened the time to negative (difference=-0.77 days, hazard ratio (HR)=1.40, 95% CI 0.97 to 2.01, p=0.072), and 120 mg/day CEP did not show the trend. Among de novo patients in the per-protocol set (PPS), 60 mg/day CEP significantly shortened the time to negative (difference=-0.87 days, HR=1.56, 95% CI 1.03 to 2.37, p=0.035). Among viral rebound patients in the mITT population, neither 120 mg/day nor 60 mg/day CEP significantly shortened the time to negative compared to placebo. Adverse events were not different among the three groups, and no serious adverse events occurred. Treatment of asymptomatic or mild Covid-19 with 120 mg/day or 60 mg/day CEP did not shorten the time to negative compared with placebo, without evident safety concerns. Among de novo infected patients with good compliance, 60 mg/day CEP significantly shortened the time to negative compared with placebo (NCT05398705).

## Introduction

Coronavirus disease 2019 (COVID-19), caused by severe acute respiratory syndrome coronavirus 2 (SARS-CoV-2), has caused over 500 million confirmed cases and places a heavy burden on global health.^1^ SARS-CoV-2 has shown a remarkably fast evolutionary rate, and five major variants of SARS-CoV-2 (Alpha, Beta, Gamma, Delta, and Omicron) have emerged to date.^2^ In view of the large number of confirmed cases of COVID-19 across rich and poor regions,^3^ the rapid evolution of SARS-CoV-2 and limited and expensive antiviral drugs, it is urgent to identify effective, widely available and inexpensive drugs with broad-spectrum antiviral ability against SARS-CoV-2.

Drug repurposing is a feasible approach in a pandemic. Cepharanthine (CEP) is a biscoclaurine alkaloid extracted from Stephania Cepharantha Hayata that has been used to treat various diseases (including leukopenia, alopecia and snake bites).^4,5^ CEP was identified as a potential SARS-CoV-2 antiviral drug via high-throughput screening in a recent interactome-informed drug repurposing study.^6^ It has been reported that CEP is capable of blocking viral entry by binding the SARS-CoV-2 S protein to interfere with the interaction of the S protein and its receptor angiotensin-converting enzyme 2 (ACE-2).^7-10^ Moreover, SARS-CoV-2 nonstructural protein 13 (Nsp13) is important for the replication of the viral genome and responsible for viral viability.^11^ CEP displays antiviral activity in inhibiting Nsp13 ATPase (helicase), thus inhibiting viral replication.^12^ CEP shows potential antiviral activities against SARS-CoV-2 in vitro and in vivo.^13,14^ In addition, CEP, a natural alkaloid that has been used for a long time in the treatment of various diseases, shows no significant side effects in patients.^15^ Given the well-established safety profile and significant unique antiviral effect of CEP, repurposing inexpensive and easily available CEP as an antiviral drug for the treatment of COVID-19 is of great potential.

However, information about the efficacy and safety of CEP in inhibiting SARS-CoV-2 replication to treat COVID-19 is still lacking, and clinical trials are urgently needed. Thus, during the epidemic of COVID-19 in Shanghai, China, we started a randomized, double-blind and placebo-controlled clinical trial to evaluate the safety and efficacy of CEP in inhibiting SARS-CoV-2. Given the high transmissibility but low pathogenicity of Omicron, the dominant SARS-CoV-2 variant at present, most of SARS-CoV-2-infected patients are asymptomatic or mild.^16^ Therefore, patients enrolled in this study were nonhospitalized adults with asymptomatic and mild COVID-19, and as the key index to evaluate the efficacy, the time from randomization to negative nasopharyngeal swab was the primary endpoint in this study.

## Results

### Patients

Between May 31 and July 24, 2022, a total of 551 patients were screened for inclusion at two sites in Shanghai, China, 262 de novo SARS-CoV-2 infected patients and 124 viral rebound patients underwent randomization, respectively. Among the randomized patients, 52 de novo infected patients and 17 viral rebound patients at Renji Hospital’s alternate care site did not receive the intervention drugs between June 1, 2022, and June 3, 2022, in addition, 22 de novo infected patients and 14 viral rebound patients were confirmed to be SARS-CoV-2 negative by nucleic acid test at randomization (see the supplementary data for detail reasons). These 105 randomized patients could not be evaluated for efficacy, therefore, the modified intention-to-treat (mITT) set rather than the intention-to-treat (ITT) set was selected for efficacy analysis in this study. Consequently, 188 de novo infected patients and 93 viral rebound patients were included in the mITT set, and 317 patients were included in the safety analysis set [Figure 1]. Among de novo infected patients in the mITT population, the number of patients in the 120 mg/day CEP, 60 mg/day CEP and placebo groups was 65, 68 and 55, respectively, and the number of patients who had good medication compliance and were included in the per protocol set (PPS) was 57, 52 and 43, respectively. Viral rebound patients in the mITT set allocated to the 120 mg/day CEP (n=29), 60 mg/day CEP (n=34) and placebo groups (n=30). The numbers of patients included in the PPS in the three groups were 29, 33 and 28, respectively. All patients in the mITT set had completed the 28-day follow-up [Figure 1].

**Figure 1.**
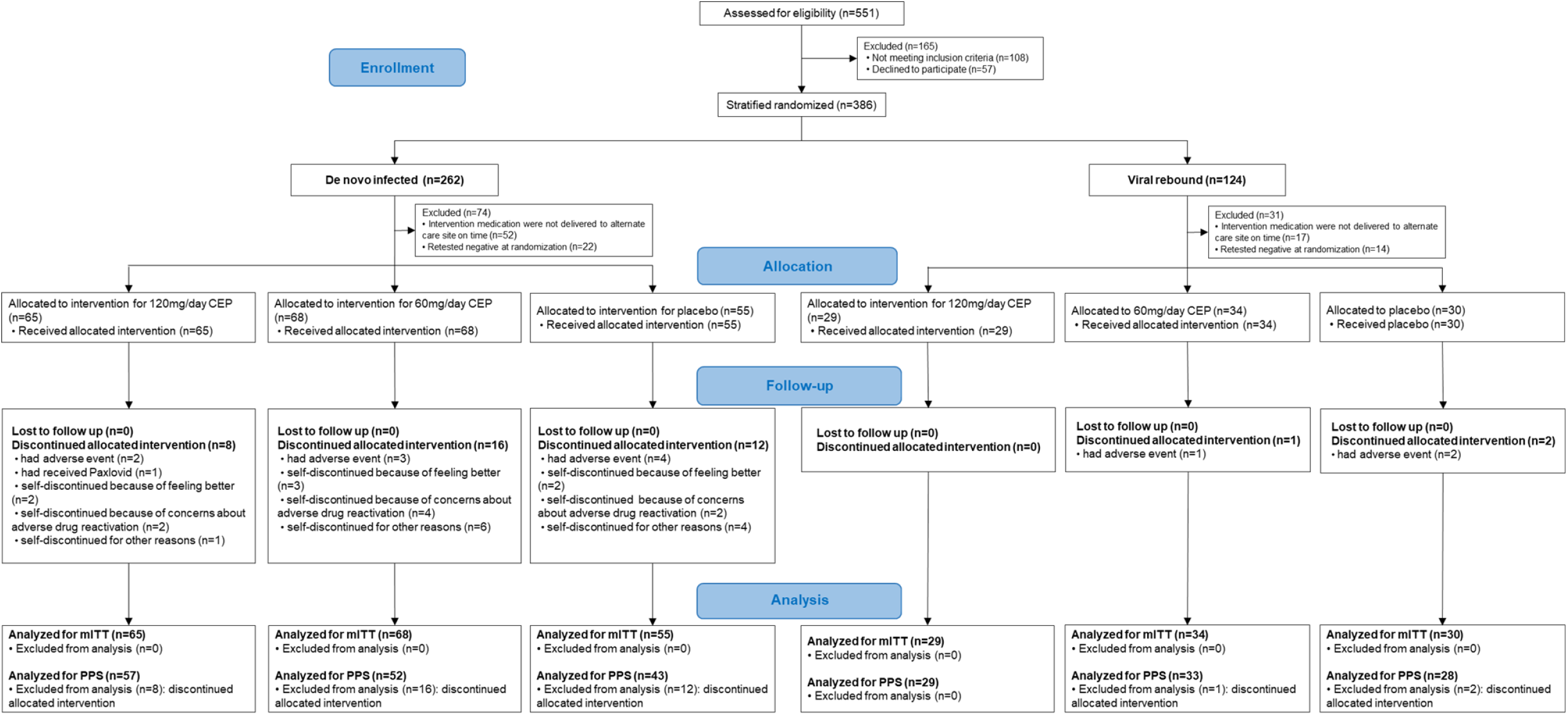
Randomization, treatment assignments, and follow-up. The figure shows that patients were recruited through May 31, 2022, to July 24, 2022, from two alternate care sites in Shanghai, China, and underwent stratified randomization according to de novo infection or viral rebound of SARS-CoV-2. Treatment assignments and follow-up were conducted.

In the mITT population, the median age of de novo infected patients in the 120 mg/day CEP, 60 mg/day CEP and placebo groups was 41.00 years [31.00, 54.00], 35.50 years [26.50, 49.25] and 43.00 years [31.50, 51.50], respectively, and the median age of viral rebound patients was 55.00 years [46.00, 62.00], 48.00 years [36.00, 55.00], and 44.50 years [35.25, 57.50], respectively. Among de novo infected patients, the proportions of patients over 60 years of age in the three groups were 13.8% (9/65), 10.3% (7/68), and 9.1% (5/55), and those among viral rebound patients were 31.0% (9/29), 8.8% (3/34), and 20.0% (6/30), respectively. The proportions of male patients in the three groups were 69.2% (45/65), 50.0% (34/68), and 63.6% (35/55) among de novo infected patients and 79.3% (23/29), 76.5% (26/34), and 66.7% (20/30) among viral rebound patients, respectively. Similar to the course features of de novo infection and viral rebound of SARS-CoV-2, patients with symptoms were mostly in the de novo infection population, in which the proportions of mild COVID-19 patients who had symptoms before enrollment in the three groups were 67.7% (44/65), 61.8% (42/68), and 60.0% (33/55), and the proportions of patients who still had symptoms at enrollment were 29.2% (19/65), 23.5% (16/68) and 30.9% (17/55), respectively. Among viral rebound patients, the proportions of mild COVID-19 patients who had symptoms before enrollment in the three groups were 10.3% (3/29), 11.8% (4/34), and 6.7% (2/30), of whom 3.4% (1/29), 8.8% (3/34) and 3.3% (1/30) still had symptoms upon enrollment, respectively. The percentages of de novo infected patients who were at high risk of developing severe COVID-19 were 53.8% (35/65), 53.7% (36/68) and 49.1% (27/55) in the three groups, of which the percentages of patients with chronic underlying diseases were 20.0% (13/65), 20.6% (14/68) and 16.4% (9/55), respectively. Among viral rebound patients, high-risk patients accounted for 62.1% (18/29), 73.5% (25/34) and 66.7% (20/30) of the three groups, of which 27.6% (8/29), 23.5% (8/34) and 33.3% (10/30) had chronic underlying diseases [Table 1]. See Table 1 for other detailed characteristics of the mITT population, de novo infected patients and viral rebound patients.

**Table 1.**
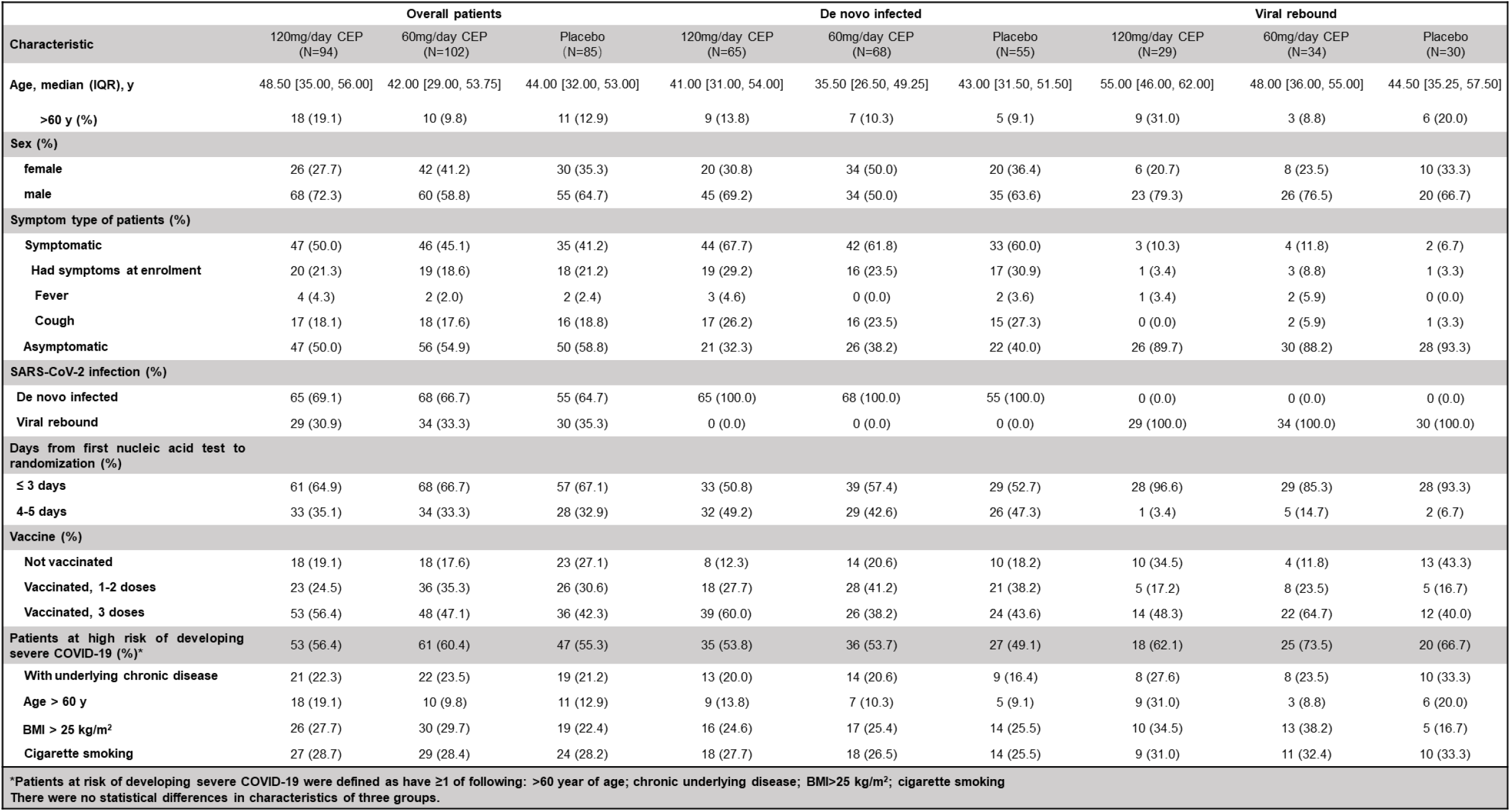
Demographic and Clinical Characteristics of the Patients (modified intention-to-treat population)

**Figure 3. Subgroup analysis of the difference in the time to negative nasopharyngeal swab compared to placebo (mITT).**
|  |  |  |  |  |  |  |  |  |  |
| --- | --- | --- | --- | --- | --- | --- | --- | --- | --- |
|  | **Overall patients** |  |  | **De novo infected** |  |  | **Viral rebound** |  |  |
| **Characteristic** | **120mg/day CEP**<br>**(N=94)** | **60mg/day CEP**<br>**(N=102)** | **Placebo**<br>**(N=85)** | **120mg/day CEP**<br>**(N=65)** | **60mg/day CEP**<br>**(N=68)** | **Placebo**<br>**(N=55)** | **120mg/day CEP**<br>**(N=29)** | **60mg/day CEP**<br>**(N=34)** | **Placebo**<br>**(N=30)** |
| **Age, median (IQR), y** | 48.50 [35.00, 56.00] | 42.00 [29.00, 53.75] | 44.00 [32.00, 53.00] | 41.00 [31.00, 54.00] | 35.50 [26.50, 49.25] | 43.00 [31.50, 51.50] | 55.00 [46.00, 62.00] | 48.00 [36.00, 55.00] | 44.50 [35.25, 57.50] |
| **>60 y (%)** | 18 (19.1) | 10 (9.8) | 11 (12.9) | 9 (13.8) | 7 (10.3) | 5 (9.1) | 9 (31.0) | 3 (8.8) | 6 (20.0) |
| **Sex (%)** |  |  |  |  |  |  |  |  |  |
| **female** | 26 (27.7) | 42 (41.2) | 30 (35.3) | 20 (30.8) | 34 (50.0) | 20 (36.4) | 6 (20.7) | 8 (23.5) | 10 (33.3) |
| **male** | 68 (72.3) | 60 (58.8) | 55 (64.7) | 45 (69.2) | 34 (50.0) | 35 (63.6) | 23 (79.3) | 26 (76.5) | 20 (66.7) |
| **Symptom type of patients (%)** |  |  |  |  |  |  |  |  |  |
| **Symptomatic** | 47 (50.0) | 46 (45.1) | 35 (41.2) | 44 (67.7) | 42 (61.8) | 33 (60.0) | 3 (10.3) | 4 (11.8) | 2 (6.7) |
| **Had symptoms at enrolment** | 20 (21.3) | 19 (18.6) | 18 (21.2) | 19 (29.2) | 16 (23.5) | 17 (30.9) | 1 (3.4) | 3 (8.8) | 1 (3.3) |
| **Fever** | 4 (4.3) | 2 (2.0) | 2 (2.4) | 3 (4.6) | 0 (0.0) | 2 (3.6) | 1 (3.4) | 2 (5.9) | 0 (0.0) |
| **Cough** | 17 (18.1) | 18 (17.6) | 16 (18.8) | 17 (26.2) | 16 (23.5) | 15 (27.3) | 0 (0.0) | 2 (5.9) | 1 (3.3) |
| **Asymptomatic** | 47 (50.0) | 56 (54.9) | 50 (58.8) | 21 (32.3) | 26 (38.2) | 22 (40.0) | 26 (89.7) | 30 (88.2) | 28 (93.3) |
| **SARS-CoV-2 infection (%)** |  |  |  |  |  |  |  |  |  |
| **De novo infected** | 65 (69.1) | 68 (66.7) | 55 (64.7) | 65 (100.0) | 68 (100.0) | 55 (100.0) | 0 (0.0) | 0 (0.0) | 0 (0.0) |
| **Viral rebound** | 29 (30.9) | 34 (33.3) | 30 (35.3) | 0 (0.0) | 0 (0.0) | 0 (0.0) | 29 (100.0) | 34 (100.0) | 30 (100.0) |
| **Days from first nucleic acid test to randomization (%)** |  |  |  |  |  |  |  |  |  |
| **≤ 3 days** | 61 (64.9) | 68 (66.7) | 57 (67.1) | 33 (50.8) | 39 (57.4) | 29 (52.7) | 28 (96.6) | 29 (85.3) | 28 (93.3) |
| **4-5 days** | 33 (35.1) | 34 (33.3) | 28 (32.9) | 32 (49.2) | 29 (42.6) | 26 (47.3) | 1 (3.4) | 5 (14.7) | 2 (6.7) |
| **Vaccine (%)** |  |  |  |  |  |  |  |  |  |
| **Not vaccinated** | 18 (19.1) | 18 (17.6) | 23 (27.1) | 8 (12.3) | 14 (20.6) | 10 (18.2) | 10 (34.5) | 4 (11.8) | 13 (43.3) |
| **Vaccinated, 1-2 doses** | 23 (24.5) | 36 (35.3) | 26 (30.6) | 18 (27.7) | 28 (41.2) | 21 (38.2) | 5 (17.2) | 8 (23.5) | 5 (16.7) |
| **Vaccinated, 3 doses** | 53 (56.4) | 48 (47.1) | 36 (42.3) | 39 (60.0) | 26 (38.2) | 24 (43.6) | 14 (48.3) | 22 (64.7) | 12 (40.0) |
| **Patients at high risk of developing severe COVID-19 (%)<sup>a</sup>** | 53 (56.4) | 61 (60.4) | 47 (55.3) | 35 (53.8) | 36 (53.7) | 27 (49.1) | 18 (62.1) | 25 (73.5) | 20 (66.7) |
| **With underlying chronic disease** | 21 (22.3) | 22 (23.5) | 19 (21.2) | 13 (20.0) | 14 (20.6) | 9 (16.4) | 8 (27.6) | 8 (23.5) | 10 (33.3) |
| **Age > 60 y** | 18 (19.1) | 10 (9.8) | 11 (12.9) | 9 (13.8) | 7 (10.3) | 5 (9.1) | 9 (31.0) | 3 (8.8) | 6 (20.0) |
| **BMI > 25 kg/m<sup>2</sup>** | 26 (27.7) | 30 (29.7) | 19 (22.4) | 16 (24.6) | 17 (25.4) | 14 (25.5) | 10 (34.5) | 13 (38.2) | 5 (16.7) |
| **Cigarette smoking** | 27 (28.7) | 29 (28.4) | 24 (28.2) | 18 (27.7) | 18 (26.5) | 14 (25.5) | 9 (31.0) | 11 (32.4) | 10 (33.3) |
| <sup>a</sup>Patients at risk of developing severe COVID-19 were defined as have ≥1 of following: >60 year of age; chronic underlying disease; BMI>25 kg/m<sup>2</sup>; cigarette smoking<br>There were no statistical differences in characteristics of three groups. |  |  |  |  |  |  |  |  |  |

### Time from randomization to negative nasopharyngeal swab

#### De novo infected patients

Among de novo infected patients in the mITT set, the time to negative nasopharyngeal swab in the 120 mg/day CEP, 60 mg/day CEP and placebo groups was 5.71 (95% CI 5.00 to 6.42), 5.01 (95% CI 4.37 to 5.66), and 5.78 (95% CI 4.88 to 6.41) days, respectively. Compared with placebo, there was no significant difference in the time to negative nasopharyngeal swab in the 120 mg/day CEP group (RMST difference=-0.07 days, HR=1.10, 95% CI 0.76 to 1.60, p=0.606), while there was a trend of a shorter time to negative nasopharyngeal swab in the 60 mg/day CEP group (RMST difference=-0.77, HR=1.40, 95% CI 0.97 to 2.01, p=0.072) [Table 2] [Figure 2].

**Table 2.** Time from randomization to negative nasopharyngeal swab in three groups.

|  | Overall patients |  |  |  |  |  | De novo infected patients |  |  |  |  |  | Viral rebound patients |  |  |  |  |  |
| --- | --- | --- | --- | --- | --- | --- | --- | --- | --- | --- | --- | --- | --- | --- | --- | --- | --- | --- |
|  | mITT |  |  | PPS |  |  | mITT |  |  | PPS |  |  | mITT |  |  | PPS |  |  |
|  | 120mg/day CEP | 60mg/day CEP | Placebo | 120mg/day CEP | 60mg/day CEP | Placebo | 120mg/day CEP | 60mg/day CEP | Placebo | 120mg/day CEP | 60mg/day CEP | Placebo | 120mg/day CEP | 60mg/day CEP | Placebo | 120mg/day CEP | 60mg/day CEP | Placebo |
| **N (missing)** | 94 (0) | 102 (0) | 85 (0) | 86 (0) | 85 (0) | 71 (0) | 65 (0) | 68 (0) | 55 (0) | 57 (0) | 52 (0) | 43 (0) | 29 (0) | 34 (0) | 30 (0) | 29 (0) | 33 (0) | 28 (0) |
| **Outcome event (%)** | 94 (100) | 102 (100) | 85 (100) | 86 (100) | 85 (100) | 71 (100) | 65 (100) | 68 (100) | 55 (100) | 57 (100) | 52 (100) | 43 (100) | 29 (100) | 34 (100) | 30 (100) | 29 (100) | 33 (100) | 28 (100) |
| **Time to negative\* (95%CI), days** | 4.70<br>(4.11, 5.30) | 4.15<br>(3.65, 4.65) | 4.58<br>(3.89, 5.26) | 4.56<br>(3.93, 5.18) | 3.89<br>(3.37, 4.42) | 4.35<br>(3.61, 5.10) | 5.71<br>(5.00, 6.42) | 5.01<br>(4.37, 5.66) | 5.78<br>(4.88, 6.68) | 5.63<br>(4.86, 6.41) | 4.85<br>(4.12, 5.58) | 5.72<br>(4.69, 6.75) | 2.45<br>(1.97, 2.93) | 2.41<br>(2.13, 2.70) | 2.37<br>(2.07, 2.67) | 2.45<br>(1.96, 2.93) | 2.39<br>(2.10, 2.68) | 2.25<br>(2.06, 2.44) |
| **Difference (95%CI)** | 0.13<br>(-0.78, 1.03) | -0.43<br>(-1.28, 0.42) | - | 0.21<br>(-0.77, 1.18) | -0.46<br>(-1.37, 0.45) | - | -0.07<br>(-1.12, 1.07) | -0.77<br>(-1.88, 0.34) | - | -0.09<br>(-1.38, 1.20) | -0.87<br>(-2.14, 0.39) | - | 0.08<br>(-0.49, 0.65) | 0.05<br>(-0.37, 0.46) | - | 0.20<br>(-0.32, 0.72) | 0.14<br>(-0.20, 0.49) | - |
| **Hazard Ratio (95%CI)** | 1.07<br>(0.78, 1.45) | 1.21<br>(0.91, 1.63) | - | 1.00<br>(0.72, 1.39) | 1.16<br>(0.84, 1.60) | - | 1.10<br>(0.76, 1.60) | 1.40<br>(0.97, 2.01) | - | 1.11<br>(0.73, 1.68) | 1.56<br>(1.03, 2.37) | - | 1.02<br>(0.59, 1.77) | 0.85<br>(0.51, 1.42) | - | 0.97<br>(0.56, 1.70) | 0.79<br>(0.46, 1.4) | - |
| **P value\*\*** | 0.683 | 0.193 | - | 0.788 | 0.439 | - | 0.606 | 0.072 | - | 0.619 | 0.035 | - | 0.941 | 0.534 | - | 0.928 | 0.382 | - |

\*Time to negative was the time from randomization to negative nasopharyngeal swab, it was evaluated by RMST.
\*\*p value was evaluated by Cox proportional hazards model, adjusted for de novo or viral rebound, underlying chronic disease, age (>60 yr or ≤60 yr), gender, symptoms, days from first nucleic acid test to randomization .

**Figure 2.**
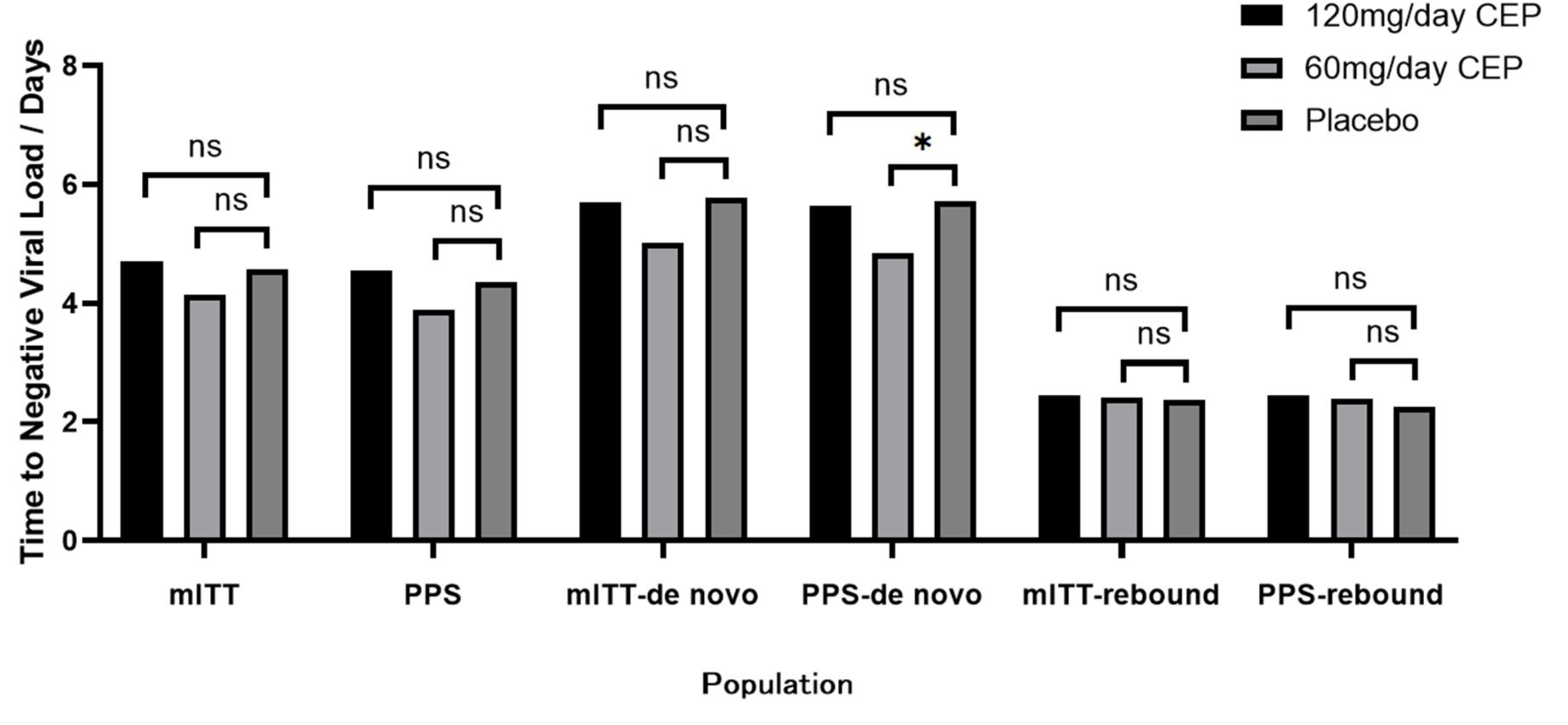
Time to negative viral load in three groups of different populations. The bar plot shows the primary endpoint of time from randomization to negative nasopharyngeal swab; from left to right are the times of the 120 mg/day CEP, 60 mg/day CEP and placebo groups in the mITT set, patients with good medication compliance (PPS), de novo infection patients in the mITT set, de novo infection patients in the PPS, viral rebound patients in the mITT set and viral rebound patients in the PPS. * indicates a significant difference between two groups, ns indicates no significant difference.

**Figure 3.**
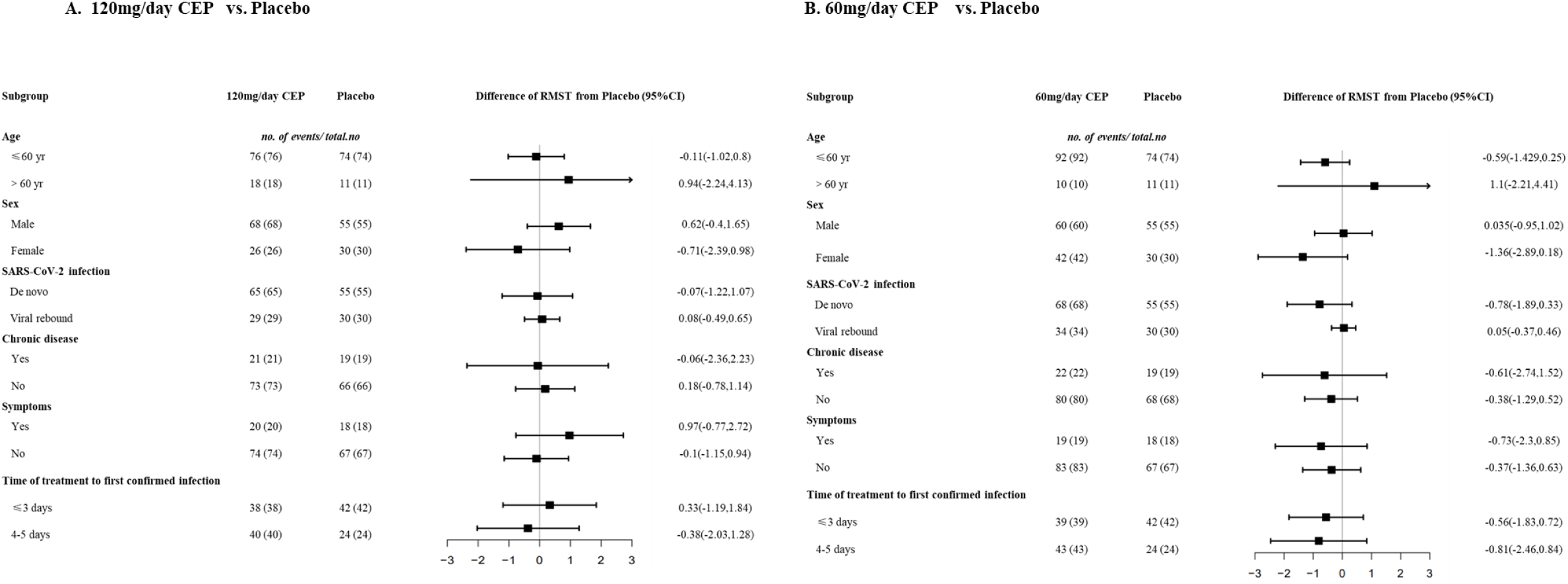
Subgroup analysis of the difference in the time to negative nasopharyngeal swab compared to placebo (mITT). Panel A shows the subgroup analysis of the difference in the time from randomization to negative nasopharyngeal swab between patients who received 120 mg/day of CEP and those who received placebo. Panel B shows the subgroup analysis of the difference in the time from randomization to negative nasopharyngeal swab between patients who received 60 mg/day of CEP and those who received placebo. The differences in the time were estimated by RMST. There was no significant difference in any subgroup.

Among de novo infected patients who had good medication compliance (PPS), the time to negative nasopharyngeal swab in the 60 mg/day CEP and placebo groups was 4.85 (95% CI 4.12 to 5.58) and 5.72 (95% CI 4.69 to 5.75), respectively; 60 mg/day of CEP significantly shortened the time by 0.87 days compared with placebo (RMST difference= -0.87, HR=1.56, 95% CI 1.03 to 2.37, p=0.035), while 120 mg/day did not show efficacy (RMST difference=-0.09, HR=1.11, 95% CI 0.73 to 1.68, p=0.619) [Table 2] [Figure 2].

In the subgroup analysis among de novo infected patients in the mITT set, we observed a benefit with 60 mg/day of CEP compared with placebo among patients who were not vaccinated (RMST difference = -3.43 days, HR=3.36, 95% CI 1.04 to 10.82) and female patients (RMST difference = -2.37 days, HR=2.29, 95% CI 1.17 to 4.47) [Table S3].

#### Viral rebound patients

Among viral rebound patients in the mITT set, the time to negative nasopharyngeal swab in the 120 mg/day CEP, 60 mg/day CEP and placebo groups was 2.45 (95% CI 1.97 to 2.93), 2.41 (95% CI 2.13 to 2.70) and 2.37 (95% CI 2.07 to 2.67) days, respectively. Compared with placebo, neither 120 mg/day of CEP nor 60 mg/day of CEP significantly shortened the time. Further analysis in the PPS also showed that there was no significant difference in the time to negative swab between the CEP treatment groups and the placebo group [Table 2] [Figure 2].

#### Total population

In the mITT population, the time from randomization to negative nasopharyngeal swab as the primary outcome in the 120 mg/day CEP, 60 mg/day CEP and placebo groups was 4.7 (95% CI 4.11 to 5.30), 4.15 (95% CI 3.65 to 4.65) and 4.58 (95% CI 3.89 to 5.26) days, respectively. Compared with the placebo group, neither CEP group showed a significant efficacy in shortening the time to negative nasopharyngeal swab (120 mg/day CEP: RMST difference=0.13 days, HR=1.07, 95% CI 0.78 to 1.45, p=0.683; 60 mg/day CEP: RMST difference= -0.43 days, HR=1.21, 95% CI 0.91 to 1.63, p=0.193) [Table 2] [Figure 2].

#### Secondary outcome

None of the patients with de novo infection or viral rebound in the mITT set progressed to severe COVID-19 during the 28-day follow-up.

Among de novo infected patients, the proportions of patients who were SARS-CoV-2 positive after negative nasopharyngeal swabs in the 120 mg/day CEP, 60 mg/day CEP and placebo groups were 3.1% (2/65), 1.5% (1/68) and 5.5% (3/55), respectively. Among viral rebound patients, the proportions of patients who were positive in the three groups were 3.4% (1/29), 0.0% (0/34) and 3.3% (1/30), respectively [Table S2].

#### Safety

A total of 317 patients were included in the safety analysis set, and we compared safety endpoints among the 120 mg/day CEP, 60 mg/day CEP and placebo groups. There were 156 adverse events that emerged during the treatment period. The incidence rates of adverse events that emerged during or after the treatment period in the 120 mg/day CEP, 60 mg/day CEP and placebo groups were 33.65% (35/104), 37.72% (43/114) and 35.35% (35/99), respectively. Adverse events were similar in the three groups and included diarrhea, increased bowel movement frequency, drowsiness, dizziness, night sweats and others. No serious adverse events were reported in any safety analysis population. There were 81 adverse events considered by the site investigator to be related to the trial drug, and the incidence rates in the three groups were 24.04% (25/104), 23.68% (27/114) and 20.20% (20/99). The most frequently reported events (that affected at least 1% of patients) were diarrhea (13 (12.50%) in the 120 mg/day CEP group vs. 9 (7.89%) in the 60 mg/day CEP group vs. 9 (9.09%) in the placebo group), drowsiness (6 (5.77%) in the 120 mg/day CEP group vs. 10 (8.77%) in the 60 mg/day CEP group vs. 7 (7.07%) in the placebo group), night sweats (6 (5.77%) in the 120 mg/day CEP group vs. 9 (7.89%) in the 60 mg/day CEP group vs. 1 (1.01%) in the placebo group), and dizziness (3 (2.88%) in the 120 mg/day CEP group vs. 0 (0%) in the 60 mg/day CEP group vs. 1 (1.01%) in the placebo group). During the treatment period, there were no grade 3 or 4 adverse events, and all events were mild to moderate (grade 1 or 2) and were resolved at the time of this analysis [Table 3].

**Table 3.** Summary of Adverse Events, Serious Adverse Events and Adverse Events Leading to Discontinuation through Day 28 (Safety Analysis Population*)

|  |  |  |  |
| --- | --- | --- | --- |
| **Adverse Event Category** | **120mg/day CEP**<br>**(N=104)** | **60mg/day CEP**<br>**(N=114)** | **Placebo**<br>**(N=99)** |
| **Events that emerged during treatment period** |  |  |  |
| No. of adverse events | 48 | 66 | 42 |
| **Patients with adverse events — no. (%)** |  |  |  |
| Any adverse event | 35(33.65) | 43(37.72) | 35(35.35) |
| Serious adverse event | 0(0) | 0(0) | 0(0) |
| Maximum grade 3 or 4 adverse event | 0(0) | 0(0) | 0(0) |
| Maximum grade 5 adverse event | 0(0) | 0(0) | 0(0) |
| Discontinued drug or placebo because of adverse event | 2(1.92) | 4(3.51) | 6(6.06) |
| Had dose reduction or temporary discontinuation owing to adverse event | 1(0.96) | 1(0.88) | 1(1.01) |
| **Adverse Event Type — no. (%)** |  |  |  |
| Diarrhea | 19 (18.27) | 22 (19.30) | 15 (15.15) |
| Increased bowel frequency | 9(8.65) | 15 (13.16) | 10 (10.10) |
| Drowsiness | 6 (5.77) | 10 (8.77) | 7 (7.07) |
| Dizzy | 3 (2.88) | 0 (0) | 1 (1.01) |
| Night sweats | 6 (5.77) | 9 (7.89) | 1 (1.01) |
| Others | 5 (4.81) | 10 (8.77) | 8 (8.08) |
| **Events considered to be related to drug or placebo** |  |  |  |
| No. of adverse events | 29 | 31 | 21 |
| **Patients with adverse events — no. (%)** |  |  |  |
| Any adverse event | 25(24.04) | 27(23.68) | 20(20.20) |
| Serious adverse event | 0(0) | 0(0) | 0(0) |
| Maximum grade 3 or 4 adverse event | 0(0) | 0(0) | 0(0) |
| Maximum grade 5 adverse event | 0(0) | 0(0) | 0(0) |
| Discontinued drug or placebo because of adverse event | 2(1.92) | 3(2.63) | 4(4.04) |
| Had dose reduction or temporary discontinuation owing to adverse event | 0(0) | 1(0.88) | 1(1.01) |
| **Adverse Event Type — no. (%)** |  |  |  |
| Diarrhea | 13 (12.50) | 9 (7.89) | 9 (9.09) |
| Drowsiness | 6 (5.77) | 10 (8.77) | 7 (7.07) |
| Night sweats | 6 (5.77) | 9 (7.89) | 1 (1.01) |
| Dizzy | 3 (2.88) | 0 (0) | 1 (1.01) |
| Others | 1 (0.96) | 3 (2.63) | 3 (3.03) |
| * Shown are data for all patients who received at least one dose of drug or placebo. |  |  |  |

## Discussion

This study is the first randomized placebo-controlled trial to evaluate the safety and efficacy of oral administration of CEP for asymptomatic or mild COVID-19 patients at alternate care sites in China. We explored the safety and efficacy of viral clearance of two different doses of CEP in treating patients with de novo infection and viral rebound of SARS-CoV-2. The results showed that the side effects of both 5-day oral administration of 60 mg/day of CEP and 120 mg/day of CEP were generally mild and without safety concerns. Among de novo infected patients in the mITT set, 60 mg/day of CEP showed a tendency to shorten the time to negative swab, and among de novo infected patients with good medication compliance (PPS), 60 mg/day of CEP significantly accelerated viral clearance compared with placebo. However, 120 mg/day of CEP did not shorten the time to negative swabs.

Among de novo SARS-CoV-2-infected patients, although 60 mg/day of CEP significantly shortened the time to negative swabs in the PPS (HR=1.56, 95% CI 1.03 to 2.37, p=0.035), it did not achieve a significant difference in the mITT set (HR=1.40, 95% CI 0.97 to 2.01, p=0.072). Therefore, the efficacy of 60 mg/day of CEP in the PPS should be carefully explained. The possible reason may be the limited sample size. The number of de novo infected patients who underwent randomization was 262, which did not reach the designated sample size of 315 patients, and this study may not have had enough power to detect a significant difference. Based on the hazard ratio of this study, a sufficient sample size may be required for further validation trials.

As a natural alkaloid with multiple targets^5^, CEP has anti-SARS-CoV-2, antioxidant (scavenging free radicals), cell membrane stabilization, drug efflux transporter inhibition, vasodilatation and other properties. However, the specific dose of CEP required to activate each target and the interactions between the targets are not completely clear. In this study, when the dose of CEP was increased to 120 mg/day, it did not show an efficacy in shortening the time to negative nasopharyngeal swab among de novo infected patients (neither in the mITT set nor in the PPS). Clearance of viruses in patients is determined by the speed of virus replication and the speed of the immune system in eliminating the virus. The possible reason is that in addition to the direct antiviral effect against SARS-CoV-2 ^13,14^, 120 mg/day of CEP may inhibit the ability of the immune system to clear SARS-CoV-2. It has been reported that 10 mg/kg of CEP in mice can not only inhibit the replication of SARS-CoV-2^14^ but also inhibit the NF-κB pathway and reduce the release of cytokines such as TNF-α, IL-1β and IL-6.^17^ In addition, CEP inhibits neutrophils^18^, inhibits dendritic cells^19^, stimulates T cells^20^, etc. According to this study, 60 mg/day of CEP significantly shortened the time to negative results among de novo infected patients with good compliance. We speculated that at a dose of 60 mg/day, CEP did not activate the target of inhibiting the immune system, while a dose of 120 mg/day of CEP could activate the target. As a result, 120 mg/day of CEP did not shorten the time to negative results.

It has been reported that the viral shedding time of de novo SARS-CoV-2-infected and viral rebound patients is quite different^21^. Therefore, we designed a stratified randomization of de novo infection and viral rebound of SARS-CoV-2. The results of our study also support those of previous reports: in the mITT set, the average time to negative nasopharyngeal swab of patients with viral rebound was 2.42 days, which was 2.3 times shorter than the 5.5 days for de novo infected patients. Neither 60 mg/day nor 120 mg/day of CEP shortened the time to negativity among SARS-CoV-2 viral rebound patients. The first possible reason is that some viral rebound patients in this study may have been false-positive for SARS-CoV-2 infection, which may have led to bias in evaluating the time to negative results. It has been reported that among patients with SARS-CoV-2 viral rebound, live viruses cannot be isolated, and most of these viruses are RNA fragments of SARS-CoV-2 detected by PCR^21^, which indicates that these viral rebound patients may be mixed with patients who are not truly infected (false-positive SARS-CoV-2 infection). However, in this real-world clinical trial at alternate care sites, we were unable to identify these false-positive patients effectively. Second, the limited sample size may account for the results that did not show any significant difference.

As a natural alkaloid, CEP has been used to treat various diseases for a long time and has been proven to have good safety,^4,5^ which was also shown in the treatment of asymptomatic and mild COVID-19. In this study, no serious adverse events occurred in the 120 mg/day CEP, 60 mg/day CEP or placebo group. The incidence of adverse events in the three groups was similar, and the most common adverse events were diarrhea and drowsiness.

This trial has several strengths. This study is the first randomized clinical trial in the world, to our knowledge, to evaluate the safety and efficacy of oral administration of CEP for nonhospitalized adults with asymptomatic or mild COVID-19. Second, this study was designed as a randomized, double-blind, placebo-controlled trial and had high quality, ensuring that the results were scientific. Third, patients selected for this study were nonhospitalized adult patients with asymptomatic or mild COVID-19, which means that the results can be extrapolated. At present, because the Omicron variant has low pathogenicity but high infectivity^2^, asymptomatic and mild infections are the most common and will continue to be so in the future. Therefore, the results of this study can be better applied to the overwhelming majority of SARS-CoV-2-infected people.

There are also some limitations of this study. First, the primary analysis population was the mITT population rather than the intention-to-treat (ITT) population in this study. Sixty-nine patients did not receive the intervention medicine, and 36 patients had negative nasopharyngeal swabs upon retesting at randomization. Therefore, we chose the mITT population as the primary analysis population in our study. The reason that these 69 patients were excluded from the analysis was that random numbers were generated, but the intervention medicines were not received; rather, these patients refused to take the first dose of medicine for subjective reasons. Therefore, we consider that the exclusion of these 69 patients (consecutively excluded from June 3 to June 5) had little impact on the randomization. Although a total of 105 patients were excluded after randomization, there was no significant difference in characteristics at baseline in the mITT population, de novo infected subgroup or viral rebound subgroup, which also indicated that the exclusion of these patients was not destructive to randomization. In future validation studies, to avoid this problem, measures should be taken to ensure that all enrolled participants receive the intervention and are confirmed to be positive for SARS-CoV-2 by PCR at randomization. Second, since no patients developed pneumonia or severe COVID-19 in this study, the effect of CEP on preventing severe COVID-19 could not be evaluated. This is reasonable for the characteristics of Omicron variant infection. The Omicron variant is the major epidemic variant of SARS-CoV-2, and its pathogenicity is lower than that of other known variants, which leads to most of SARS-CoV-2-infected patients being asymptomatic or mild^16^, and the rate of developing severe COVID-19 is extremely low^22^. Third, patients enrolled in this study were limited to asymptomatic or mild COVID-19 patients, and the efficacy of CEP for severe COVID-19 is unknown and needs further study. However, according to the epidemic characteristics of the majority of asymptomatic or mild SARS-CoV-2-infected patients, the number of patients enrolled in this study was appropriate.

In conclusion, these results provide evidence that oral administration of 120 mg/day of CEP and 60 mg/day of CEP for 5 days is safe for asymptomatic or mild COVID-19 patients. Neither 120 mg/day nor 60 mg/day of CEP shortened the time to negative nasopharyngeal swab among the de novo infection or viral rebound SARS-CoV-2 patients. However, 60 mg/day of CEP significantly shortened the time to viral shedding among de novo SARS-CoV-2-infected patients with good compliance. In future studies, the efficacy of CEP in treating COVID-19 may be validated among de novo SARS-CoV-2-infected patients at a dose of 60 mg/day.

## Methods

### Study design and patients

This was a double-blind, stratified randomized, parallel, placebo-controlled trial. Participants were recruited between May 31, 2022 and July 24, 2022, from the alternate care site at Shanghai New International Expo Centre, China (managed by Renji Hospital, School of Medicine, Shanghai Jiaotong University) and alternate care site at Shanghai Chongming Fuxing, China (managed by Xinhua Hospital, School of Medicine, Shanghai Jiaotong University).

The inclusion criteria for this study included age from 16 to 85 years, confirmed SARS-CoV-2 infection by PCR, SARS-CoV-2 infection for less than 5 days prior to randomization, diagnosis of asymptomatic or mild COVID-19 according to WHO guidelines^1^, and signed informed consent. Key exclusion criteria included pneumonia or severe COVID-19^23^, acute exacerbation of chronic underlying diseases, and pregnancy or lactation. Asymptomatic COVID-19 was defined as a positive SARS-CoV-2 test without symptoms^24^. Mild COVID-19 was defined as confirmed COVID-19 infection complicated with mild symptoms, including fever, cough, or changes in taste or smell, without evidence of dyspnea or pneumonia on imaging^24^. Patients at high risk of developing severe COVID-19 were required to have ≥1 of the following characteristics or comorbidities associated with an increased risk of developing severe COVID-19 illness: ≥60 years of age, body mass index (BMI) >25 kg/m2, cigarette smoking, and chronic underlying disease (including diabetes, hypertension, chronic lung, cardiovascular, kidney, or immunosuppressive disease, or other medically complex conditions)^25-27^.

Considering that viral rebound patients have a lower viral load and faster viral conversion to negativity than de novo SARS-CoV-2 infection patients, we designed a stratified randomization based on SARS-CoV-2 de novo infection or viral rebound. The enrolled patients were classified into de novo SARS-CoV-2 infection and viral rebound groups. De novo SARS-CoV-2 infection was defined as SARS-CoV-2 infection for the first time (positive nasopharyngeal swab test by PCR: Ct ≤35 for the ORF1ab or nucleocapsid N genes). Viral rebound was defined as retested SARS-CoV-2 positivity after negative conversion of previous SARS-COV-2 infection. The negative conversion of SARS-CoV-2 was defined as two consecutive negative nasopharyngeal swabs (Ct value>35 for the ORF1ab and N genes) of SARS-CoV-2 at least 24 hours apart^28,29^. The trial was reviewed and approved by the Renji Hospital Ethics Committee, School of Medicine, Shanghai Jiaotong University (KY2022-107-A) and the Xinhua Hospital Ethics Committee, School of Medicine, Shanghai Jiaotong University (XHEC-C-2022-064-1). All participants provided written informed consent. This trial was registered on ClinicalTrials.gov, NCT05398705.

### Randomization and masking

Stratified randomization was performed according to de novo SARS-CoV-2 infection and viral rebound. A centralized randomization system provided by the Clinical Information Management Suite (CIMS) Medical Technology Company (Chengdu, China) combined with a drug dispensation system provided by Alibaba Health Information Technology Limited were used to realize the randomization and masking. The CIMS was responsible for generating a randomization number, and Alibaba Health Information Technology Limited was responsible for linking the randomization number to the drug and placebo and removing the manufacturer’s label for masking. The three groups were blinded with 12 patients as the number of each block. Researchers, patients, caregivers and statisticians were masked to allocation, and a separate unblinded data monitoring committee evaluated safety throughout this study. All personnel were unblinded following premature study termination until all patients had completed follow-up, at which point the study continued in an unblinded fashion.

### Procedures

This study enrolled patients from May 31, 2022, to July 24, 2022, at two alternate care sites in Shanghai, China: 1) from May 31, 2022, to June 11, 2022, at the alternate care site of the Shanghai New International Expo Centre and 2) from July 16 to July 24, 2022 at the alternate care site of Shanghai Chongming Fuxing. Eligible mITT patients were stratified randomly into de novo infection and viral rebound groups. Randomized patients were divided 1:1:1 by means of a centralized randomization system to receive 40 mg of CEP, 20 mg of CEP or matched placebo orally every 8 hours for 5 days (15 doses total) or until negative conversion. CEP and matching placebo were manufactured by Yun Nan Bai Yao Pharmaceutical Group Inc. (Z20026797). All patients at alternate care sites received standard medical treatment according to the Scheme for Diagnosis and Treatment of 2019 Novel Coronavirus Pneumonia (The 9^th^ Trial Edition) from the Health Commission of China, including bed rest, adequate energy and nutrition, plenty of water, traditional Chinese medicine treatment, etc^30^. At the alternate care sites, nasopharyngeal swabs were collected for nucleic acid testing, and the results were recorded. Adverse events and development of pneumonia or severe COVID-19 were recorded during the alternate care site visits and follow-up (until Day 28 after randomization). Patients were followed up by telemedicine visits every week after leaving the alternate care site.

### Outcomes

The primary outcome of efficacy of viral clearance was the time from randomization to negative nasopharyngeal swab, which was defined as the duration from randomization to negative conversion (the first of two consecutive negative nasopharyngeal swabs tested by PCR, Ct value>35 for the ORF1ab and N genes). The secondary outcomes were the proportion of patients who progressed to pneumonia or severe COVID-19 and the proportion of patients who were SARS-CoV-2 positive after a negative nasopharyngeal swab.

Safety endpoints included adverse events, serious adverse events, and adverse events that contributed to discontinuation of the study intervention that occurred during the treatment period and during the follow-up period. Reported adverse events were coded according to the Medical Dictionary for Regulatory Activities (MedDRA), version 25.0. The safety analysis population included all patients who received at least one dose of the study intervention drug. The incidence data for each treatment group were analyzed within the safety analysis population.

### Statistical analysis

All eligible patients who received at least 1 dose of the drug or placebo who also tested positive by nucleic acid test at randomization were included in the mITT population, with patients who completed the treatment (had good medical compliance (80%-120%)) included in the PPS. All patients who received at least 1 dose of the drug or placebo were included in the safety analysis set (SS).

In the original design of this study, the time from randomization to negative nasopharyngeal swab was used as the primary endpoint. The sample size of this study was determined to detect a potential clinical superiority of CEP in time to viral shedding. A total of 105 patients in each group would provide 80% power to detect a hazard ratio of 1.5, which was tested in the Cox proportional hazards model of the time to negative nasopharyngeal swab for CEP over placebo. The overall probability of an event was 0.9, with a 2-sided significance level of α=0.05, and the ratio of the sample in each group was 1:1:1. Considering the explorative property of this study and that the assumptions in sample size determination were based on limited clinical evidence, this study continued to enroll patients after 315 patients were enrolled when research resources were sufficient.

The analysis for efficacy assessment was performed in both the mITT set and the PPS. The analysis performed in the mITT set was primary analysis, and that performed in the PPS was considered supportive analysis. Continuous variables are presented as the means with standard deviations or medians with interquartile ranges (IQRs), and categorical variables are reported as numbers and percentages. There were no missing data in the mITT set.

Since all patients were quarantined until they were negative for SARS-CoV-2 infection, there was no censoring for time to negative nasopharyngeal swab due to loss to follow-up in this study. Death was treated as a competitive event of viral shedding. The time to negative nasopharyngeal swab of each group was summarized, and differences between groups were assessed using the log-rank test. A Cox proportional hazards model was fitted to estimate the HR and 95% confidence intervals for the 60 mg/day CEP group and 120 mg/day CEP group compared with the control group, and the reported p value was evaluated by the Cox proportional hazards model. In this model, SARS-CoV-2 infection (de novo or viral rebound), underlying disease (presence or absence), age (>60 years or ≤60 years), sex (male or female), symptoms on admission (symptomatic or asymptomatic) and days from first nucleic acid test to randomization were included as adjusted variables. In addition, the restricted mean survival time (RMST) and corresponding 95% CI of each group were also estimated. Prespecified subgroup analyses of primary and secondary endpoints were conducted, and 95% CIs were provided to evaluate whether the treatment effect varied according to age, sex, symptoms (asymptomatic or mild), SARS-CoV-2 infection (de novo or viral rebound), or high-risk factors for progression to severe COVID-19 (including age≥60 years, smoking, obesity, and underlying clinical conditions)^25-27^.

## Data Availability

All data produced in the present study are available upon reasonable request to the authors
All data produced in the present work are contained in the manuscript
All data produced are available online at clinical trials.gov or send an email to

## Acknowledgments

This work was supported by the Shanghai Municipal Education Commission-Gaofeng Clinical Medicine Grant Support (20152213). Cepharanthine and placebo were supplied by Yun Nan Bai Yao Pharmaceutical Group Inc. (Z20026797) without charge. We are grateful to all the patients who participated in this study and their families. We thank BLUEBALLON Beijing (China) and the study teams for monitoring and management.

## Author contributions

Jianyi Wei^#1^ and Shuming Pan^#2^ contributed equally to this work, and share joint first authorship. Hai Li and Chen Yao are joint corresponding authors. Concept and design: Hai Li, Chen Yao, Biyun Qian, Ning Zhang, Qimin Zhan. Statistical analysis: Chen Yao, Yongpei Yu, Lanlan Yu, Weituo Zhang. Original drafting of the manuscript: Jianyi Wei, Shupeng Liu, Yan Zhang, Zixuan Shen. Reviewing and editing draft for important intellectual content: Hai Li, Chen Yao, Junhua Zheng, Weituo Zhang, Yongpei Yu. Administrative, technical and material support: Hai Li, Shumimng Pan, Chen Yao, Ning Zhang, Qiming Zhan, Jun Wang. Data collection: Jianyi Wei, Shupeng Liu, Zixuan Shen, Yuexiang Bian, Adila·ABuduaini, Fuchen Dong, Xin Zhang, Jinhui Li, Wei Zhai, Qixiang Song, Yu Zheng, Lei Li and Weihua Pan. All authors had full access to all the data in the study and had final responsibility for the decision to submit for publication.

---

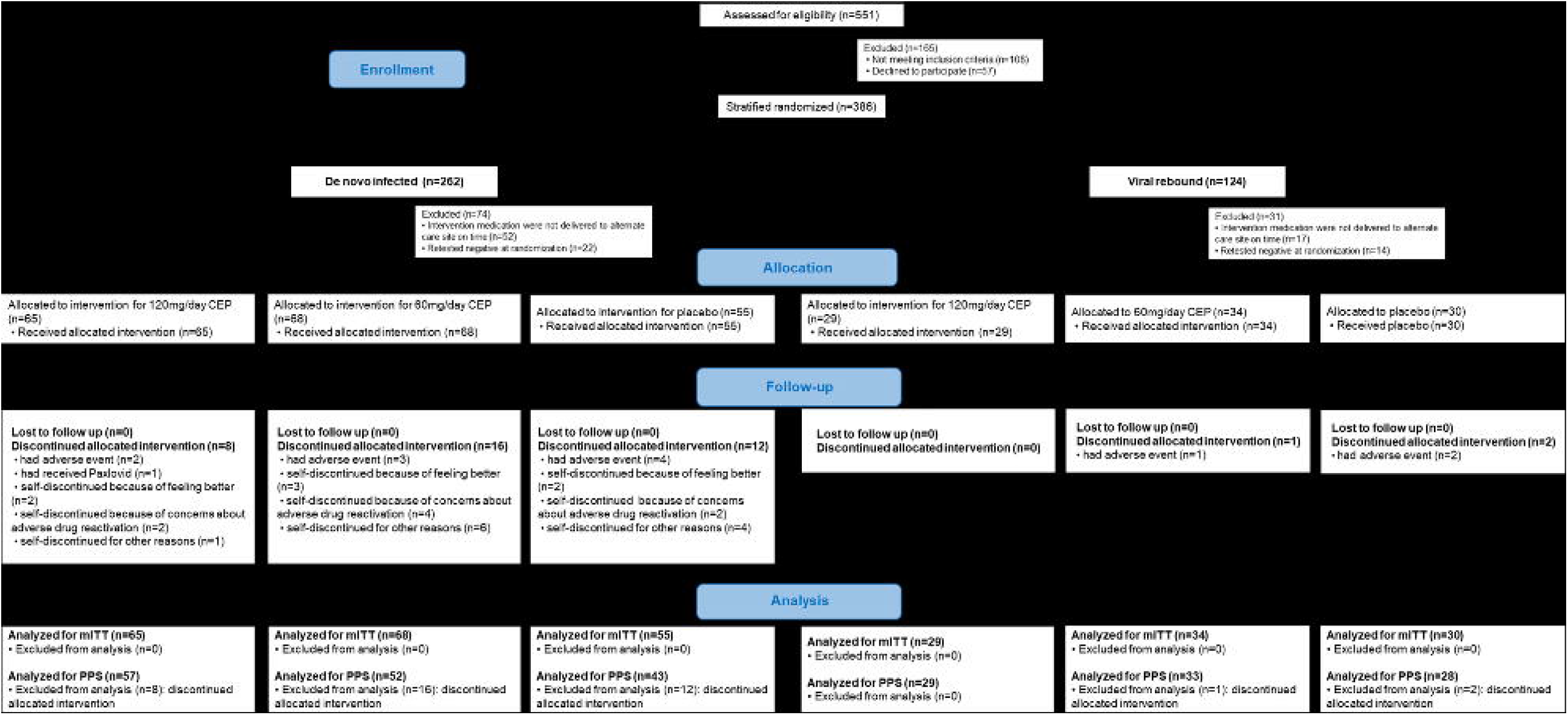

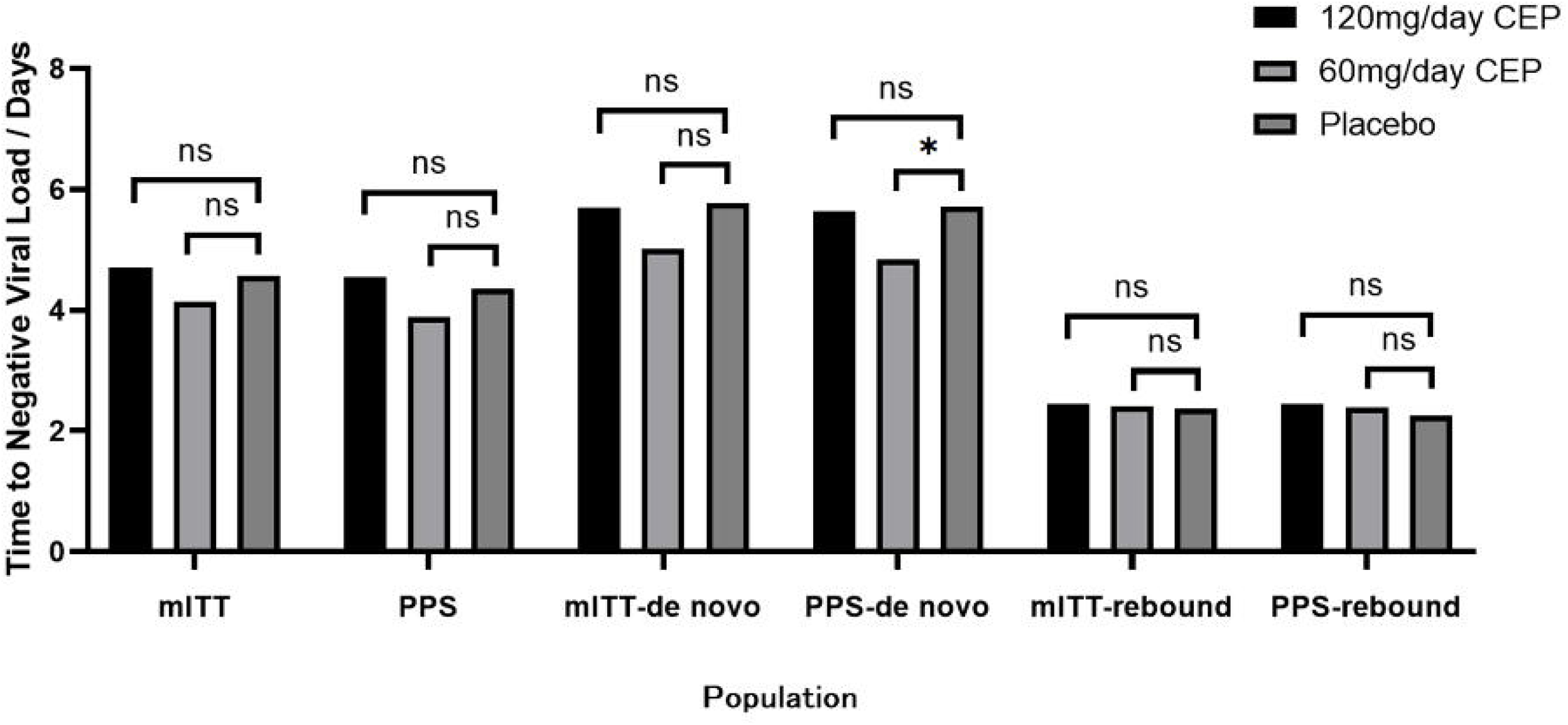

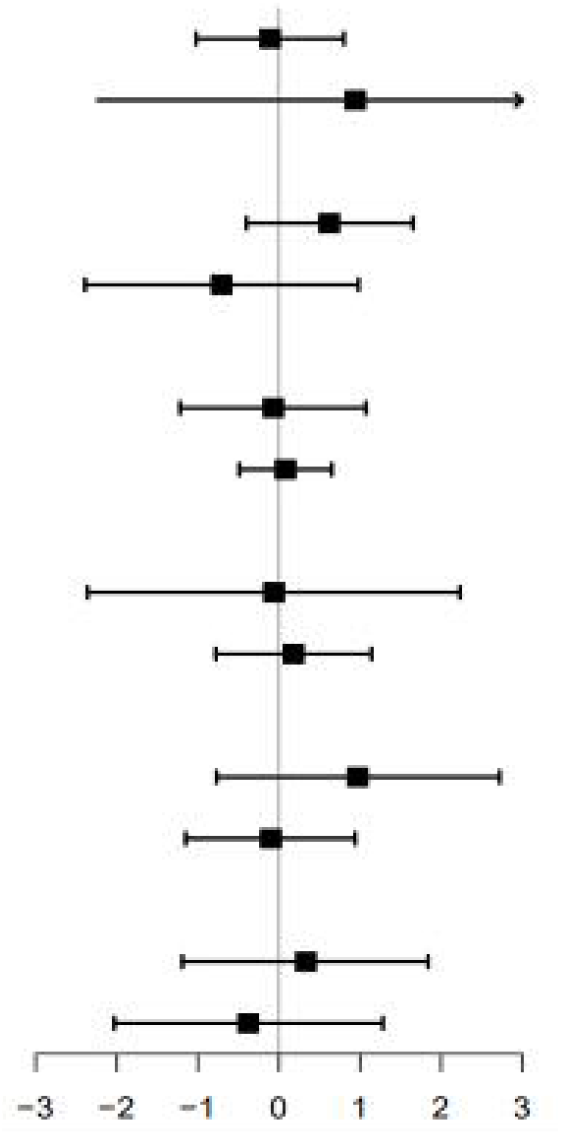

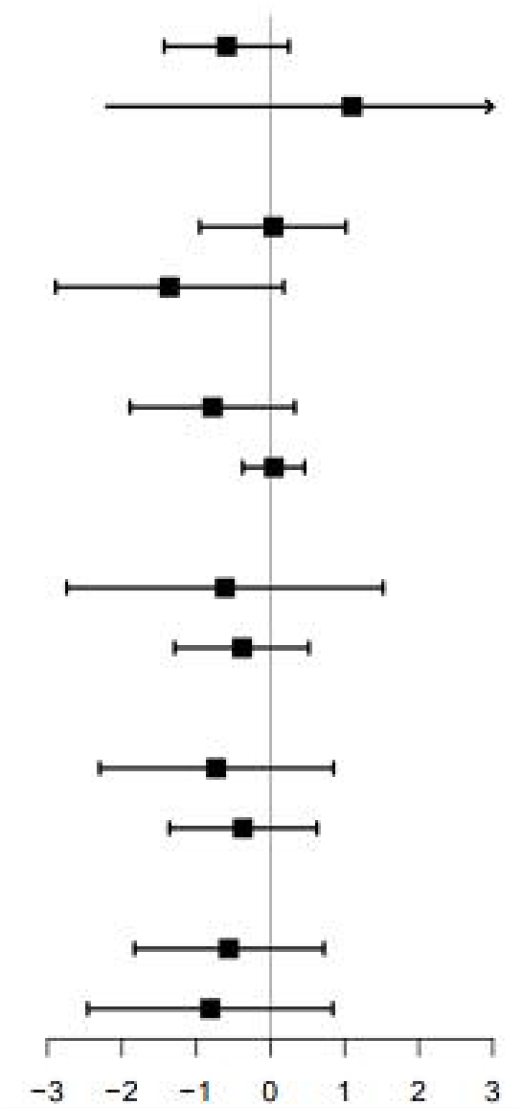

## Reference

1. WHO coronavirus (COVID-19) dashboard. World Health Organization, https://covid19.who.int (2022)

2. Jacobs JL, Haidar G, Mellors JW. COVID-19: Challenges of Viral Variants. Annu Rev Med. https://www.annualreviews.org/doi/10.1146/annurev-med-042921-020956 (2022)

3. COVID-19 Cumulative Infection Collaborators. Estimating global, regional, and national daily and cumulative infections with SARS-CoV-2 through Nov 14, 2021: a statistical analysis. Lancet 399(10344):2351-2380 (2022).

4. Rogosnitzky M, Okediji P, Koman I. Cepharanthine: a review of the antiviral potential of a Japanese-approved alopecia drug in COVID-19. Pharmacol Rep 72(6):1509–1516 (2020).

5. Bailly C. Cepharanthine: An update of its mode of action, pharmacological properties and medical applications. Phytomedicine 62:152956 (2019).

6. Riva L, Yuan S, Yin X, et al. Discovery of SARS-CoV-2 antiviral drugs through large-scale compound repurposing. Nature 586(7827):113–119 (2020).

7. Ayele AG, Enyew EF, Kifle ZD. Roles of existing drug and drug targets for COVID-19 management. Metabol Open 11:100103 (2021).

8. Sixto-López Y, et al. Structural insights into SARS-CoV-2 spike protein and its natural mutants found in Mexican population. Sci Rep 11(1):4659 (2021).

9. Ohashi H, Watashi K, Saso W, et al. Potential anti-COVID-19 agents, cepharanthine and nelfinavir, and their usage for combination treatment. iScience 24(4):102367 (2021).

10. Lan J, Ge J, Yu J, et al. Structure of the SARS-CoV-2 spike receptor-binding domain bound to the ACE2 receptor. Nature 581(7807):215–220 (2020).

11. White MA, Lin W, Cheng X. Discovery of COVID-19 Inhibitors Targeting the SARS-CoV-2 Nsp13 Helicase. J Phys Chem Lett 11(21):9144–9151 (2020).

12. Hossain R, Sarkar C, Hassan SMH, et al. In Silico Screening of Natural Products as Potential Inhibitors of SARS-CoV-2 Using Molecular Docking Simulation. Chin J Integr Med 28(3):249–256 (2022).

13. Fan HH, Wang LQ, Liu WL, et al. Repurposing of clinically approved drugs for treatment of coronavirus disease 2019 in a 2019-novel coronavirus-related coronavirus model. Chin Med J (Engl) 133(9):1051–1056 (2020).

14. Zhang S, Huang W, Ren L, et al. Comparison of viral RNA-host protein interactomes across pathogenic RNA viruses informs rapid antiviral drug discovery for SARS-CoV-2. Cell Res 32(1):9–23 (2022).

15. Rogosnitzky M, Danks R. Therapeutic potential of the biscoclaurine alkaloid, cepharanthine, for a range of clinical conditions. Pharmacol Rep 63(2):337–47 (2011).

16. Epidemic distribution of novel coronavirus pneumonia. Chinese Center for Disease Control and Prevention. https://2019ncov.chinacdc.cn/2019-nCoV/index.html (2022)

17. Samra YA, et al. Cepharanthine and Piperine ameliorate diabetic nephropathy in rats: role of NF-κB and NLRP3 inflammasome. Life Sci 157:187–199 (2016).

18. Murakami K, Okajima K, Uchiba M. The prevention of lipopolysaccharide-induced pulmonary vascular injury by pretreatment with cepharanthine in rats. Am J Respir Crit Care Med 161(1):57–63 (2000).

19. Uto T, Nishi Y, Toyama M, Yoshinaga K, Baba M. Inhibitory effect of cepharanthine on dendritic cell activation and function. Int Immunopharmacol 11(11):1932–8 (2011).

20. Xu W, et al. Molecular mechanisms and therapeutic implications of tetrandrine and cepharanthine in T cell acute lymphoblastic leukemia and autoimmune diseases. Pharmacol Ther 217:107659 (2021).

21. Lu Li JT, et al. Characteristics of SARS-CoV-2 Delta variant-infected individuals with intermittently positive retest viral RNA after discharge. National Science Review 9(10):nwac141 (2022).

22. Fan Y, Li X, Zhang L, Wan S, Zhang L, Zhou F. SARS-CoV-2 Omicron variant: recent progress and future perspectives. Signal Transduct Target Ther 7(1):141 (2022).

23. Berlin DA, Gulick RM, Martinez FJ. Severe Covid-19. N Engl J Med 383(25):2451–2460 (2020).

24. Gandhi RT, Lynch JB, Del Rio C. Mild or Moderate Covid-19. N Engl J Med 383(18):1757–1766 (2020).

25. Docherty AB, et al. Features of 20 133 UK patients in hospital with covid-19 using the ISARIC WHO Clinical Characterisation Protocol: prospective observational cohort study. BMJ 369:m1985 (2020).

26. Zhou F, et al. Clinical course and risk factors for mortality of adult inpatients with COVID-19 in Wuhan, China: a retrospective cohort study. Lancet 395(10229):1054–1062 (2020).

27. Chen T, et al. Clinical characteristics of 113 deceased patients with coronavirus disease 2019: retrospective study. BMJ 368:m1091 (2020).

28. Dighe A, et al. Response to COVID-19 in South Korea and implications for lifting stringent interventions. BMC Med 18(1):321 (2020).

29. Xu Y, et al. Characteristics of pediatric SARS-CoV-2 infection and potential evidence for persistent fecal viral shedding. Nat Med 26(4):502–505 (2020).

30. National Health Commission of the People’s Republic of China. Chinese guideline for the management covid-19 (version 9.0, in Chinese). http://www.nhc.gov.cn/yzygj/s7653p/202203/b74ade1ba4494583805a3d2e40093d88/files/ef09aa4 070244620b010951b088b8a27.pdf (2022)

