## Supplementary materials for "Safety and Efficacy of Oral administrated Cepharanthine in Non-hospitalized, asymptomatic or mild COVID-19 patients: A Double-blind, Randomized, Placebo-controlled Trial"

### Methods

##### Detailed reasons for patient exclusion from mITT analysis

Because the alternate care site in China was a quarantine site, before entering this area, the investigators first communicated with patients by telephone and obtained informed consent, determined the patients to be eligible for enrollment and performed randomization. Then, the investigator in charge of medicine distribution entered the alternate care site to distribute the medicine to the patients. From June 1, 2022, to June 3, 2022, the investigator in charge of medicine distribution had close contact with the COVID-19 patients because the working time at the alternate care site exceeded the specified requirements and was quarantined for three days according to the epidemic prevention policy in China. Therefore, between June 1, 2022, and June 3, 2022, 69 patients underwent randomization but did not receive the intervention medicine. In addition, among the randomized patients, 36 patients were confirmed to be SARS-CoV-2 negative by PCR at randomization.

##### Cepharanthine and Placebo Information

The chemical formula of CEP is $C_{37}H_{38}N_2O_6$, the molecular weight is 606.71, The CEP tablets in the study were manufactured by Yun Nan Bai Yao pharmaceutical Group Inc (Z20026797), the appearance of CEP tablet is a white or yellowish, smelled odorless and tasted bitter. One CEP tablet contains 20mg CEP.

The placebo is supplied by Yun Nan Bai Yao pharmaceutical Group Inc. The placebo is a cepharathine-matched placebo, the appearance is a milky white or yellowish tablet with appropriate hardness and wear resistance which has no obvious difference from the appearance of CEP tablets.

CEP and placebo should be stored in a normal temperature. All packaging will be labeled to indicate that the product is for investigational use. CEP and placebo were distributed by investigators in alternate care sites.

##### Dosing and Administration

120mg/day CEP group: 2 CEP tablets (40mg) orally each time, three times daily for 5 days or until SARS-CoV-2 turned consecutive negative tested by PCR.

60mg/day CEP group: 1 CEP tablet (20mg) plus 1 placebo tablet orally each time, three times daily for 5 days or until SARS-CoV-2 turned consecutive negative tested by PCR.

Placebo group: 2 placebo tablets orally each time, three times daily for 5 days or until SARS-CoV-2 turned consecutive negative tested by PCR.

##### Participant Monitoring

The daily and follow-up assessments were monitored, and sites were actively notified of events requiring review, including serious adverse events. During the alternate care sites period, participants were visited everyday for assessments including any adverse events, occurrence or disappearance of symptoms, temperature and the development of pneumonia or severe covid-19. After leaving the alternate care site, participants were followed up by telephone visit every week, until day 28 after randomization. Adverse events, viral rebound and development of Covid-19 were recorded.

##### Ethical Conduct

All patients provided written informed consent. The study was conducted in accordance with consensus ethical principles derived from international guidelines including the Declaration of Helsinki, Council for International Organizations of Medical Sciences International Ethical Guidelines, International Council for Harmonisation Good Clinical Practice guidelines, and applicable laws and regulations. The protocol and related documents were approved by an institutional review board/ethics committee before study commencement.

**Study Responsibilities**

Researcher took responsibility for study design and conduct; data collection, and interpretation; and writing of this manuscript. Faculty of Statistics, Clinical Research Center, Peking University, assisted with study design, data collection and cleaning, and statistical analyses. Data were recorded by clinical research coordinators. Clinical research coordinators and Clinical Information Management Suite(CIMS) database personnel reviewed the collected data. The clinical research coordinators then entered confirmed data into the Clinical Information Management Suite database for statistical analyses performed by and reviewed by the senior statistician in accordance with Good Clinical Practice guidelines. Cepharanthine and matching placebo were manufactured by Yun Nan Bai Yao company, and blinding of the tablets was performed by Yun Nan Bai Yao company through over-encapsulation. All the data were available to all the authors, who vouch for the accuracy and completeness of the data as well as the adherence of the trial to the protocol.

| Table S1. Demographic and Clinical Characteristics of the Per-protocol Set (PPS). | | | | | | | | | |
|---|---|---|---|---|---|---|---|---|---|
| | Overall patients | | | De novo infected | | | Viral rebound | | |
| Characteristic | 120mg/day CEP (N=86) | 60mg/day CEP (N=85) | Placebo (N=71) | 120mg/day CEP (N=57) | 60mg/day CEP (N=52) | Placebo (N=43) | 120mg/day CEP (N=29) | 60mg/day CEP (N=33) | Placebo (N=28) |
| Age, median (IQR), y | 49.00 [35.25, 56.00] | 45.00 [29.00, 55.00] | 45.00 [33.50, 54.00] | 41.00 [34.00, 54.00] | 41.00 [28.50, 53.75] | 46.00 [32.50, 53.00] | 55.00 [46.00, 62.00] | 48.00 [36.00, 55.00] | 44.50 [34.50, 56.50] |
| >60 y (%) | 17 (19.8) | 10 (11.8) | 10 (14.1) | 8 (14.0) | 7 (13.5) | 5 (11.6) | 9 (31.0) | 3 (9.1) | 5 (17.9) |
| Sex (%) | | | | | | | | | |
| female | 22 (25.6) | 32 (37.6) | 24 (33.8) | 16 (28.1) | 25 (48.1) | 15 (34.9) | 6 (20.7) | 7 (21.2) | 9 (32.1) |
| male | 64 (74.4) | 53 (62.4) | 47 (66.2) | 41 (71.9) | 27 (51.9) | 28 (65.1) | 23 (79.3) | 26 (78.8) | 19 (67.9) |
| Symptom type of patients (%) | | | | | | | | | |
| Symptomatic | 41 (47.7) | 37 (43.5) | 25 (35.2) | 38 (66.7) | 33 (63.5) | 24 (55.8) | 3 (10.3) | 4 (12.1) | 1 (3.6) |
| Had symptoms at enrolment | 19 (22.1) | 17 (20.0) | 15 (21.1) | 18 (31.6) | 14 (26.9) | 14 (32.6) | 1 (3.4) | 3 (9.1) | 1 (3.6) |
| Fever | 4 (4.7) | 2 (2.4) | 0 (0.0) | 3 (5.3) | 0 (0.0) | 0 (0.0) | 1 (3.4) | 2 (6.1) | 0 (0.0) |
| Cough | 16 (18.6) | 16 (18.8) | 15 (21.1) | 16 (28.1) | 14 (26.9) | 14 (32.6) | 0 (0.0) | 2 (6.1) | 1 (3.6) |
| Asymptomatic | 45 (52.3) | 48 (56.5) | 46 (64.8) | 19 (33.3) | 19 (36.5) | 19 (44.2) | 26 (89.7) | 29 (87.9) | 27 (96.4) |
| SARS-CoV-2 infection (%) | | | | | | | | | |
| De novo infected | 57 (66.3) | 52 (61.2) | 43 (60.6) | 57 (100.0) | 52 (100.0) | 43 (100.0) | 0 (0.0) | 0 (0.0) | 0 (0.0) |
| Viral rebound | 29 (33.7) | 33 (38.8) | 28 (39.4) | 0 (0.0) | 0 (0.0) | 0 (0.0) | 29 (100.0) | 33 (100.0) | 28 (100.0) |
| Days from first nucleic acid test to randomization (%) | | | | | | | | | |
| ≤ 3 days | 58 (67.4) | 60 (70.6) | 47 (66.2) | 30 (52.6) | 32 (61.5) | 21 (48.8) | 28 (96.6) | 28 (84.8) | 26 (92.9) |
| 4-5 days | 28 (32.6) | 25 (29.4) | 24 (33.8) | 27 (47.4) | 20 (38.5) | 22 (51.2) | 1 (3.4) | 5 (15.2) | 2 (7.1) |
| Vaccine (%) | | | | | | | | | |
| Not vaccinated | 17 (19.8) | 13 (15.3) | 20 (28.2) | 7 (12.3) | 10 (19.2) | 8 (18.6) | 10 (34.5) | 3 (9.1) | 12 (42.9) |
| Vaccinated, 1-2 doses | 20 (23.2) | 30 (35.3) | 19 (26.8) | 15 (26.3) | 22 (42.3) | 14 (32.6) | 5 (17.2) | 8 (24.2) | 5 (17.9) |
| Vaccinated, 3 doses | 49 (57.0) | 42 (49.4) | 32 (45.1) | 35 (61.4) | 20 (38.5) | 21 (48.8) | 14 (48.3) | 22 (66.7) | 11 (39.3) |
| Patients at high risk of developing severe COVID-19 (%)** | 48 (55.8) | 54 (63.5) | 41 (57.7) | 30 (52.6) | 30 (57.7) | 22 (51.2) | 18 (62.1) | 24 (72.7) | 19 (67.9) |
| With underlying chronic disease | 20 (23.3) | 22 (25.9) | 17 (23.9) | 12 (21.1) | 12 (23.1) | 8 (18.6) | 8 (27.6) | 10 (30.3) | 9 (32.1) |
| Age > 60 y | 17 (19.8) | 10 (11.8) | 10 (14.1) | 8 (14.0) | 7 (13.5) | 5 (11.6) | 9 (31.0) | 3 (9.1) | 5 (17.9) |
| BMI > 25 kg/m² | 24 (27.9) | 23 (27.1) | 16 (22.5) | 14 (24.6) | 11 (21.2) | 11 (25.6) | 10 (34.5) | 12 (36.4) | 5 (17.9) |
| Cigarette smoking | 23 (26.7) | 28 (32.9) | 20 (28.2) | 14 (24.6) | 17 (32.7) | 11 (25.6) | 9 (31.0) | 11 (33.3) | 9 (32.1) |

*Shown are data for all patients who underwent randomization, were not retested negative at randomization and received at least one dose of drug or placebo.
**Patients at risk of developing severe COVID-19 were defined as have ≥1 of following: >60 year of age; chronic underlying disease; BMI>25 kg/m²; cigarette smoking
There were no statistical differences in characteristics of three groups.

| Table S2. Secondary outcomes in three groups of mITT population. (28 Days) | | | | |
|---|---|---|---|---|
| | 60 mg/day CEP (N=102) | 120 mg/day CEP (N=94) | Placebo (N=85) | P value |
| Patients developed to pneumonia or severe COVID-19 — no.(%) | 0(0) | 0(0) | 0(0) | - |
| SARS-CoV-2 returned to positive after turning negative — no.(%) | 1 (1.0) | 3 (3.2) | 4 (4.7) | 0.258 |
| Hospitalization for COVID-19 — no.(%) | 0(0) | 0(0) | 0(0) | - |

**Table S3. Subgroup analysis of the differences in the time from randomization to negative nasopharyngeal swab in the 60 mg/day CEP and placebo groups among de novo infected patients. (mITT)**

| Subgroup | No. of events | | Time to negative swab | | RMST difference (95% CI) | Hzard Ratio (95% CI) | P value |
|---|---|---|---|---|---|---|---|
| | 60 mg/day CEP | Placebo | 60 mg/day CEP | Placebo | | | |
| Age | | | | | | | |
| ≤ 60yr | 61 | 50 | 4.75（4.10,5.41） | 5.54（4.69,6.39） | -0.79 (-1.86,0.29) | 1.35 (0.92,1.98) | 0.127 |
| > 60yr | 7 | 5 | 7.14（5.03,9.25） | 8.20（3.58,12.82） | -1.06 (-6.14,4.02) | 1.65 (0.46,5.93) | 0.442 |
| Sex | | | | | | | |
| Male | 34 | 35 | 5.32（4.38,6.26） | 5.06（4.08,6.03） | 0.27 (-1.09,1.62) | 0.93 (0.56,1.56) | 0.783 |
| Female | 34 | 20 | 4.68（3.79,5.56） | 7.05（5.40,8.70） | -2.37 (-4.25,-0.5) | 2.29 (1.17,4.47) | **0.015** |
| Symptoms | | | | | | | |
| Yes | 16 | 17 | 4.31（3.03,5.59） | 4.94（3.81,6.08） | -0.63 (-2.34,1.08) | 1.38 (0.63,3.01) | 0.417 |
| No | 52 | 38 | 5.21（4.47,5.96） | 6.16（4.98,7.34） | -0.95 (-2.34,0.45) | 1.49 (0.96,2.32) | 0.073 |
| Days from first nucleic acid test to randomization | | | | | | | |
| ≤3 days | 39 | 29 | 5.67（4.73,6.61） | 6.52（5.44,7.59） | -0.85 (-2.28,0.58) | 1.29 (0.79,2.1) | 0.307 |
| 4-5 days | 29 | 26 | 4.10（3.37,4.84） | 4.96（3.55,6.38） | -0.86 (-2.45,0.74) | 1.17 (0.66,2.08) | 0.583 |
| Vaccine | | | | | | | |
| not vaccinated | 14 | 10 | 4.57（3.76,5.38） | 8.00（5.48, 10.53） | -3.43 (-6.08,-0.78) | 3.36 (1.04,10.82) | **0.042** |
| ≥ 1 doses | 54 | 45 | 5.11 (4.32, 5.90) | 5.29 (4.40,6.17) | -0.18 (-1.36,1.01) | 1.11 (0.74,1.57) | 0.596 |
| Patients at high risk of developing severe COVID-19 | | | | | | | |
| yes | 36 | 27 | 5.25（4.41,6.10） | 5.89（4.53,7.25） | -0.64 (-2.24,0.97) | 1.24 (0.74,2.07) | 0.420 |
| no | 31 | 28 | 4.78（3.76,5.79） | 5.68（4.50,6.86） | -0.9 (-2.47,0.66) | 1.44 (0.85,2.42) | 0.177 |
| Chronic disease | | | | | | | |
| yes | 14 | 9 | 5.50（3.92,7.08） | 8.0（5.37,10.63） | -2.5 (-5.57,0.57) | 2.05 (0.77,5.47) | 0.151 |
| no | 54 | 46 | 4.87（4.17,5.57） | 5.35（4.45,6.24） | -0.48 (-1.62,0.66) | 1.3 (0.86,1.95) | 0.210 |

**Table S4. Subgroup analysis of the differences in the time from randomization to negative nasopharyngeal swab in the 120mg/day CEP and placebo groups among de novo infected patients. (mITT)**

| Subgroup | No. of events | | Time to negative swab | | RMST difference (95% CI) | Hzard Ratio (95% CI) | P value |
|---|---|---|---|---|---|---|---|
| | 120 mg/day CEP | Placebo | 120 mg/day CEP | Placebo | | | |
| Age | | | | | | | |
| ≤ 60yr | 56 | 50 | 5.23(4.52,5.94) | 5.54(4.69,6.39) | -0.31 (-1.41,0.8) | 1.06 (0.72,1.56) | 0.776 |
| > 60yr | 9 | 5 | 8.67(7.16,10.18) | 8.20(3.58,12.82) | 0.47 (-4.39,5.33) | 0.99 (0.13,7.7) | 0.991 |
| Sex | | | | | | | |
| Male | 45 | 35 | 5.69(4.80,6.58) | 5.06(4.08,6.03) | 0.632 (-0.69,1.95) | 0.87 (0.55,1.39) | 0.555 |
| Female | 20 | 20 | 5.75(4.63,6.88) | 7.05(5.40,8.70) | -1.3 (-3.3,0.7) | 1.48 (0.75,2.9) | 0.260 |
| Symptoms | | | | | | | |
| Yes | 17 | 19 | 5.84(4.44,7.24) | 4.94(3.81,6.08) | 0.9 (-0.9,2.7) | 0.78 (0.39,1.56) | 0.488 |
| No | 46 | 38 | 5.65(4.84,6.47) | 6.16(4.98,7.34) | -0.51 (-1.94,0.93) | 1.27 (0.79,2.02) | 0.322 |
| Days from first nucleic acid test to randomization | | | | | | | |
| ≤3 days | 33 | 29 | 6.42(5.33,7.52) | 6.52(5.44,7.59) | -0.09 (-1.63,1.44) | 1.06 (0.62,1.8) | 0.843 |
| 4-5 days | 32 | 26 | 4.97(4.15,5.78) | 4.96(3.55,6.38) | 0.007 (-1.63,1.64) | 0.9 (0.53,1.54) | 0.703 |
| Vaccine | | | | | | | |
| not vaccinated | 8 | 10 | 8.00(5.86,10.14) | 8.00(5.48,10.53) | 0 (-3.31,3.31) | 1.19 (0.31,4.61) | 0.802 |
| ≥ 1 doses | 57 | 45 | 5.39(4.68,6.10) | 5.29(4.40,10.60) | 0.097 (-1.04,1.23) | 0.96 (0.64,1.44) | 0.848 |
| Patients at high risk of developing severe COVID-19 | | | | | | | |
| yes | 35 | 27 | 6.31(5.30,7.33) | 5.89(4.53,7.25) | 0.43 (-1.28,2.13) | 0.87 (0.51,1.47) | 0.604 |
| no | 30 | 28 | 5.00(4.09,5.91) | 5.68(4.50,6.86) | -0.68 (-2.17,0.82) | 1.23 (0.72,2.1) | 0.455 |
| Chronic disease | | | | | | | |
| yes | 13 | 9 | 6.31(4.46,8.16) | 8.00(5.37,10.63) | -1.69 (-4.91,1.53) | 2.7 (0.93,7.83) | 0.068 |
| no | 52 | 46 | 5.56(4.81,6.31) | 5.35(4.45,6.24) | 0.21 (-0.96,1.38) | 0.93 (0.62,1.41) | 0.733 |

### Original and Final Protocol

### Original Protocol

#### 1. Key Trial Contacts

| | |
|---|---|
| **Chief Investigator** | Professor Hai Li<br>Professor of department of gastroenterological division, Renji Hospital<br>1630 Dongfang Road, Shanghai<br>China<br> |
| **Sponsor** | Shanghai Jiao Tong University School of Medicine |
| **Funder(s)** | No |

#### 2. Background

In the anti-SARS-CoV-2 study by high-throughput compound screening at Beijing University of Chemical Technology, Cepharanthine was found to have good inhibition of Andrographis neo-coronavirus among 2406 candidate compounds, and 10uM of Cepharanthine inhibited coronavirus replication by 15,393-fold. Because of the extremely high nucleotide sequence homology between Andrographis paniculata and human SARS-CoV-2, the above invention was recently granted a patent for the invention by the China Intellectual Property Office, claiming the use of Cepharanthine for the preparation of drugs for the treatment of SARS-CoV-2 viral infectious diseases. In the study of the mechanism of CEP inhibition of SARS-CoV-2, Japanese scholars reported that CEP inhibits viral replication by binding to the spikes of SARS-CoV-2. Second, CEP has the combined efficacy of inhibiting nuclear transcription factor NF-kB activation, lipid peroxidation, NO, and cytokine production by interfering with viral replication and host inflammatory response in multiple ways.

In an in vitro pharmacodynamic study of SARS-CoV-2 inhibition by CEP, an EC50 of 0.77 and 0.1 uM of virus inhibition was observed in SARS-CoV-2-infected A549 cell line and human lung epithelial cells, respectively.

There are no published pharmacodynamic studies of CEP in mammals infected with SRAS-CoV-2.

With respect to international clinical studies of CEP for the treatment of SRS-CoV-2, in November 2021 PharmaDrug, a Canadian publicly traded pharmaceutical company, announced that an enterosoluble CEP (PD-001) completed a pre-clinical study filing communication meeting with the U.S. FDA with the goal of conducting a registered clinical study of PD-001 for the treatment of mild to moderate COVID-19.

#### 3. Rationale

The purpose of this study is to evaluate the efficacy and safety of high/low- dose cepharanthine for the Treatment of COVID-19 in asymptomatic and nonpneumonia mild adult participants with COVID-19 who do not need to be in the hospital, but in alternate care site.

#### 4. Synopsis

| | |
|---|---|
| Trial title | An Interventional Efficacy and Safety, Phase 2/3, Double-blind, 3-arm Study to Investigate Orally Administered High/Low-dose Cepharanthine Compared With Placebo in Nonhospitalized Asymptomatic or Mild Adult Participants With COVID-19 |
| Internal ref. no. (or short title) | Study of Oral High/Low-dose Cepharanthine Compared With Placebo in Non Hospitalized Adults With COVID-19 |
| Trial registration | NCT05398705 |
| Registration time | May 21, 2022 |
| Sponsor | Shanghai Jiaotong University School of Medicine |
| Funder | No |
| Clinical Phase | Phase II |
| Trial Design | Screening participants will sign the appropriate informed consent form (ICF) prior to completion of any study procedures.<br>Patients will be randomized to one of three arms, all participants will receive standardized medical treatment (SMT) according to Scheme for Diagnosis and Treatment of 2019 Novel Coronavirus Pneumonia (The 9th Trial Edition) from health commission of China, including bed rest, adequate energy and nutrition, pay attention to water and electrolyte balance to maintain a stable internal environment, closely monitor, antiviral drug and Chinese medicine treatment, etc.<br>low-dose experimental arm：cepharanthine 60mg/day + SMT |

| | |
|---|---|
| | high-dose experimental arm： cephapharanthine 120mg/day + SMT<br>placebo control arm： placebo + SMT<br>The primary outcome measure is the time to viral clearance which defined as first positive nucleic acid test to the date of the first negative test (in two consecutive point). SARS-CoV-2 viral load was detected and quantified by RT-PCR using nasopharyngeal swabs . Ct value > 35 for both ORF1ab and N gene was considered as negativity. |
| Sample Size | 300 |
| Planned Trial Period | 28 days per participant in the main study<br>Total trial period 2 month from June 1 2022 to July 31 2022 |

#### 5. Outcome measures

| Objectives | Outcome measures | Time points |
|---|---|---|
| Primary: | Primary: | Primary: |
| To compare the efficacy of high/low-dose Cepharanthine to placebo for the treatment of COVID-19 in asymptomatic and nonpneumonia mild adult participants with COVID-19 who do not need to be in the hospital, but in alternate care site. | SARS-CoV-2 viral load | Day 28 |
| To compare the efficacy of high/low-dose Cepharanthine to placebo for the treatment of COVID-19 in asymptomatic and nonpneumonia mild adult participants with COVID-19 who do not need to be in the hospital, but in | Time to viral clearance | Day 28 |

| | | |
|---|---|---|
| alternate care site. | | |
| Secondary: | Secondary: | Secondary: |
| To compare the efficacy of high/low-dose Cepharanthine to placebo for the treatment of COVID-19 in asymptomatic and nonpneumonia mild adult participants who are at increased risk of progression to severe disease. | Proportion of participants developing COVID-19 pneumonia | Day 28 |
| To compare the efficacy of high/low-dose Cepharanthine to placebo for the treatment of COVID-19 in asymptomatic and nonpneumonia mild adult participants who are at increased risk of progression to severe disease. | Proportion of participants developing severe pneumonia | Day 28 |
| To describe the safety and tolerability of high/low-dose Cepharanthine to placebo for the treatment of COVID-19 in asymptomatic and nonpneumonia mild adult participants with COVID-19 who do not need to be in the hospital, but in alternate care site. | Incidence of Adverse Events (AEs) and Serious Adverse Events (SAEs)of CEP relative to placebo. | Day 28 |
| To compare the efficacy of high/low-dose Cepharanthine to placebo for the treatment of COVID-19 in asymptomatic and nonpneumonia mild adult | Number of days from the onset of fever until the temperature drops below 37.3°C | Day 28 |

| |
|---|
| participants with COVID-19 who do not need to be in the hospital, but in alternate care site. |

#### 6. Arms and interventions

| Arms | Assigned Interventions |
|---|---|
| Experimental: Lowdose cepharanthine + standardized medical treatment<br>Drug:<br>cepharanthine | Day 1~5: 20mg,<br>Q8H X 5 days + standardized medical treatment<br>Drug: Cepharanthine<br>Low-dose: Day 1~5: 20mg, Q8H X 5 days (60mg/day)+SMT High-dose: Day 1~5: 40mg, Q8H X 5<br>days (120mg/day) +SMT<br>SMT:standardized medical treatment according to Scheme for Diagnosis and Treatment of 2019 Novel Coronavirus Pneumonia (The 9th Trial Edition) from health commission of China, including bed rest, adequate energy and nutrition, pay attention to water and electrolyte balance to maintain a stable internal environment, closely monitor, antiviral drug and Chinese medicine treatment, etc. |
| Experimental: Highdose cepharanthine + standardized medical<br>treatment<br>Drug:<br>cepharanthine | Day 1~5: 40mg,<br>Q8H X 5 days + standardized medical treatment<br>Drug: Cepharanthine<br>Low-dose: Day 1~5: 20mg, Q8H X 5 days (60mg/day)+SMT High-dose: Day 1~5: 40mg, Q8H X 5<br>days (120mg/day) +SMT<br>SMT:standardized medical treatment according to Scheme for Diagnosis and Treatment of 2019 Novel Coronavirus Pneumonia (The 9th Trial Edition) from health commission of China, including bed rest, adequate energy and nutrition, pay attention to water and electrolyte balance to maintain a stable internal environment, closely monitor, antiviral drug and Chinese medicine treatment, etc. |

| | |
|---|---|
| Placebo Comparator:<br>placebo+standardized medical treatment<br>Drug:<br>placebo | placebo Day 1~5:<br>placebo + standardized medical treatment<br>Drug: Placebo<br>Day 1~5: placebo+SMT<br>SMT:standardized medical treatment according to Scheme for Diagnosis and Treatment of 2019 Novel Coronavirus Pneumonia (The 9th Trial Edition) from health commission of China, including bed rest, adequate energy and nutrition, pay attention to water and electrolyte balance to maintain a stable internal environment, closely monitor, antiviral drug and Chinese medicine treatment, etc. |

#### 7. Criteria

Inclusion Criteria:

- aged over 16 years old with all genders
- SARS-CoV-2 positive （laboratory-confirmed reverse transcription polymerase chain reaction (RT PCR) test)
- patient or immediate adult family member agrees to participate in this study and signs an informed consent form
- with mild covid-19 symptoms
- confirmed SARS-CoV-2 infection within 5 days prior to randomization

Exclusion Criteria:

- Confirmed SARS-CoV-2 infection within > 5 days prior to randomization
- CT shows pneumonia on admission
- diagnosed as severe or critical COVID-19 before intervention
- has a history of chronic underlying disease and acute exacerbation of that underlying disease at the time of admission
- Females who are pregnant or breastfeeding

### Final Protocol

#### 2. Key Trial Contacts

| | |
|---|---|
| **Chief Investigator** | Professor Hai Li<br>Professor of department of gastroenterological division, Renji Hospital<br>1630 Dongfang Road, Shanghai<br>China<br> |
| **Sponsor** | Shanghai Jiao Tong University School of Medicine |
| **Funder(s)** | No |

#### 8. Background

In the anti-SARS-CoV-2 study by high-throughput compound screening at Beijing University of Chemical Technology, Cepharanthine was found to have good inhibition of Andrographis neo-coronavirus among 2406 candidate compounds, and 10uM of Cepharanthine inhibited coronavirus replication by 15,393-fold. Because of the extremely high nucleotide sequence homology between Andrographis paniculata and human SARS-CoV-2, the above invention was recently granted a patent for the invention by the China Intellectual Property Office, claiming the use of Cepharanthine for the preparation of drugs for the treatment of SARS-CoV-2 viral infectious diseases. In the study of the mechanism of CEP inhibition of SARS-CoV-2, Japanese scholars reported that CEP inhibits viral replication by binding to the spikes of SARS-CoV-2. Second, CEP has the combined efficacy of inhibiting nuclear transcription factor NF-kB activation, lipid peroxidation, NO, and cytokine production by interfering with viral replication and host inflammatory response in multiple ways.

In an in vitro pharmacodynamic study of SARS-CoV-2 inhibition by CEP, an EC50 of 0.77 and 0.1 uM of virus inhibition was observed in SARS-CoV-2-infected A549 cell line and human lung epithelial cells, respectively.

There are no published pharmacodynamic studies of CEP in mammals infected with SRAS-CoV-2.

With respect to international clinical studies of CEP for the treatment of SRS-CoV-2, in November 2021 PharmaDrug, a Canadian publicly traded pharmaceutical company, announced that an enterosoluble CEP (PD-001) completed a pre-clinical study filing communication meeting with the U.S. FDA with the goal of conducting a registered clinical study of PD-001 for the treatment of mild to moderate COVID-19.

#### 9. Rationale

The purpose of this study is to evaluate the efficacy and safety of high/low- dose cepharanthine for the Treatment of COVID-19 in asymptomatic and nonpneumonia mild adult participants with COVID-19 who do not need to be in the hospital, but in alternate care site.

#### 10. Synopsis

| | |
|---|---|
| Trial title | An Interventional Efficacy and Safety, Phase 2, Double-blind, 3-arm Study to Investigate Orally Administered High/Low-dose Cepharanthine Compared With Placebo in Non-hospitalized Asymptomatic or Mild Adult Participants With COVID-19 |
| Internal ref. no. (or short title) | Study of Oral High/Low-dose Cepharanthine Compared With Placebo in Non Hospitalized Adults With COVID-19 |
| Trial registration | NCT05398705 |
| Registration time | May 26, 2022 |
| Sponsor | Shanghai Jiaotong University School of Medicine |
| Funder | No |
| Clinical Phase | Phase II |
| Trial Design | Screening participants will sign the appropriate informed consent form (ICF) prior to completion of any study procedures.<br>Patients will be randomized to one of three arms, all participants will receive standardized medical treatment (SMT) according to Scheme for Diagnosis and Treatment of 2019 Novel Coronavirus Pneumonia (The 9th Trial Edition) from health commission of China, including bed rest, adequate energy and nutrition, pay attention to water and electrolyte balance to maintain a stable internal environment, closely monitor, antiviral drug and Chinese medicine treatment, etc. |

| | |
|---|---|
| | low-dose experimental arm： cepharanthine 60mg/day + SMT<br><br>high-dose experimental arm： cepharanthine 120mg/day + SMT<br><br>placebo control arm： placebo + SMT<br><br>The primary outcome measure is the time to viral clearance which defined as first positive nucleic acid test to the date of the first negative test (in two consecutive point). SARS-CoV-2 viral load was detected and quantified by RT-PCR using nasopharyngeal swabs . Ct value > 35 for both ORF1ab and N gene was considered as negativity. |
| Sample Size | 450 |
| Planned Trial Period | 28 days per participant in the main study<br>Total trial period 2 month from May 31 2022 to August 10 2022 |

#### 11. Outcome measures

| Objectives | Outcome measures | Time points |
|---|---|---|
| Primary: | Primary: | Primary: |
| To compare the efficacy of high/low-dose Cepharanthine to placebo for the treatment of COVID-19 in asymptomatic and nonpneumonia mild adult participants with COVID-19 who do not need to be in the hospital, but in alternate care site. | Time to viral clearance which defined as the time from randomization to negative nasopharyngeal swab (date of the first in two negative consecutive tests by PCR) | Day 28 |
| Secondary: | Secondary: | Secondary: |
| To compare the efficacy of high/low-dose Cepharanthine to placebo for the treatment of COVID-19 in asymptomatic and | Proportion of participants developing COVID-19 pneumonia | Day 28 |

| | | |
|---|---|---|
| nonpneumonia mild adult participants who are at increased risk of progression to severe disease. | | |
| To compare the efficacy of high/low-dose Cepharanthine to placebo for the treatment of COVID-19 in asymptomatic and nonpneumonia mild adult participants who are at increased risk of progression to severe disease. | Proportion of participants developing severe pneumonia | Day 28 |
| To describe the safety and tolerability of high/low-dose Cepharanthine to placebo for the treatment of COVID-19 in asymptomatic and nonpneumonia mild adult participants with COVID-19 who do not need to be in the hospital, but in alternate care site. | Incidence of Adverse Events (AEs) and Serious Adverse Events (SAEs)of CEP relative to placebo. | Day 28 |
| To compare the efficacy of high/low-dose Cepharanthine to placebo for the treatment of COVID-19 in asymptomatic and nonpneumonia mild adult participants with COVID-19 who do not need to be in the hospital, but in alternate care site. | Number of days from the onset of fever until the temperature drops below 37.3°C | Day 28 |

#### 12. Arms and interventions

| Arms | Assigned Interventions |
|---|---|

| | |
|---|---|
| Experimental: Low-dose cepharanthine + standardized medical treatment<br>Drug:<br>cepharanthine | Day 1~5: 20mg,<br>Q8H X 5 days + standardized medical treatment<br>Drug: Cepharanthine<br>Low-dose: Day 1~5: 20mg, Q8H X 5 days<br>(60mg/day)+SMT High-dose: Day 1~5: 40mg, Q8H X 5<br>days (120mg/day) +SMT<br>SMT:standardized medical treatment according to Scheme for Diagnosis and Treatment of 2019 Novel Coronavirus Pneumonia (The 9th Trial Edition) from health commission of China, including bed rest, adequate energy and nutrition, pay attention to water and electrolyte balance to maintain a stable internal environment, closely monitor, antiviral drug and Chinese medicine treatment, etc. |
| Experimental: High-dose cepharanthine + standardized medical treatment<br>Drug:<br>cepharanthine | Day 1~5: 40mg,<br>Q8H X 5 days + standardized medical treatment<br>Drug: Cepharanthine<br>Low-dose: Day 1~5: 20mg, Q8H X 5 days<br>(60mg/day)+SMT High-dose: Day 1~5: 40mg, Q8H X 5<br>days (120mg/day) +SMT<br>SMT:standardized medical treatment according to Scheme for Diagnosis and Treatment of 2019 Novel Coronavirus Pneumonia (The 9th Trial Edition) from health commission of China, including bed rest, adequate energy and nutrition, pay attention to water and electrolyte balance to maintain a stable internal environment, closely monitor, antiviral drug and Chinese medicine treatment, etc. |

| | |
|---|---|
| Placebo Comparator:<br>placebo+standardized medical treatment<br>Drug:<br>placebo | placebo Day 1~5:<br>placebo + standardized medical treatment<br>Drug: Placebo<br>Day 1~5: placebo+SMT<br>SMT:standardized medical treatment according to Scheme for Diagnosis and Treatment of 2019 Novel Coronavirus Pneumonia (The 9th Trial Edition) from health commission of China, including bed rest, adequate energy and nutrition, pay attention to water and electrolyte balance to maintain a stable internal environment, closely monitor, antiviral drug and Chinese medicine treatment, etc. |

#### 13. Criteria

Inclusion Criteria:

- aged over 16 years old with all genders
- SARS-CoV-2 positive （laboratory-confirmed reverse transcription polymerase chain reaction (RT PCR) test)
- patient or immediate adult family member agrees to participate in this study and signs an informed consent form
- with mild covid-19 symptoms
- confirmed SARS-CoV-2 infection within 5 days prior to randomization

Exclusion Criteria:

- Confirmed SARS-CoV-2 infection within > 5 days prior to randomization
- CT shows pneumonia on admission
- diagnosed as severe or critical COVID-19 before intervention
- has a history of chronic underlying disease and acute exacerbation of that underlying disease at the time of admission
- Females who are pregnant or breastfeeding
